# Deep immune profiling reveals early-stage and highly coordinated immune responses in mild COVID-19 patients

**DOI:** 10.1101/2021.08.31.21262713

**Authors:** Christophe M. Capelle, Séverine Cire, Olivia Domingues, Isabelle Ernens, Fanny Hedin, Aurélie Fischer, Chantal Snoeck, Wim Ammerlaan, Maria Konstantinou, Kamil Grzyb, Alex Skupin, Cara L. Carty, Christiane Hilger, Georges Gilson, Aljosa Celebic, Antonio Del Sol, Ian M. Kaplan, Fay Betsou, Tamir Abdelrahman, Antonio Cosma, Michel Vaillant, Guy Fagherazzi, Markus Ollert, Feng Q. Hefeng

## Abstract

While immunopathology has been widely studied in severe COVID-19 patients, immunoprotective factors in non-hospitalized patients have remained largely elusive. We systematically analyzed 484 peripheral immune cell signatures, various serological parameters and TCR repertoire in a longitudinal cohort of 63 mild and 15 hospitalized patients versus 14 asymptomatic and 26 control individuals. Within three days following PCR diagnosis, we observed coordinated responses of CD4 and CD8 T cells, various antigen presenting cells and antibody-secreting cells in mild, but not hospitalized COVID-19 patients. This early-stage SARS-CoV-2-specific response was predominantly characterized by substantially expanded clonotypes of CD4 and less of CD8 T cells. The early-stage responses of T cells and dendritic cells were highly predictive for later seroconversion and protective antibody levels after three weeks in mild non-hospitalized, but not in hospitalized patients. Our systemic analysis provides the first full picture and early-stage trajectory of highly coordinated immune responses in mild COVID-19 patients.

## Introduction

The current pandemic of coronavirus disease 2019 (COVID-19) caused by severe acute respiratory syndrome corona virus 2 (SARS-CoV-2) has widely affected human health and socioeconomic layers in societies worldwide. Although vaccines, a key factor in fighting against COVID-19, have been rapidly developed and vaccination rollout has been successfully implemented in many countries^1^, a full understanding of the complexity of immune responses leading to different clinical outcomes of natural SARS-CoV-2 infection still remains incomplete. The immunopathology underlying severe COVID-19 has been thoroughly studied over the last 18 months, including antibody responses, cellular immune subsets, cytokines and chemokines that were linked to characteristics and outcome of the disease^2–6^. However, with few exceptions^7^, relatively little is known about the details of the immune response in mild and asymptomatic COVID-19 patients.

Using profiling analyses of immune cell subsets, several studies have identified crucial alterations in severe COVID-19 patients as compared to hospitalized patients with moderate disease, convalescent and healthy controls. These studies, which mainly utilized flow cytometry or single-cell mRNA sequencing for deep immune cell analysis, have demonstrated a wide spectrum of abnormal immune responses to SARS-CoV-2^4,8–10^. However, asymptomatic and/or mild COVID-19 patients have only rarely been included in these studies to draw conclusions. Thus, it remains elusive whether certain immune alterations observed in severe COVID-19 patients also occur in SARS-CoV-2 PCR-positive non-hospitalized patients with asymptomatic or mild disease. It is also unclear, whether early protective immune signatures are identifiable in asymptomatic or mild non-hospitalized patients, and how such immune signatures might compare to more severe hospitalized COVID-19 patients and healthy subjects. Overall, the connection between the orchestration of the early immune responses after natural infection with SARS-CoV-2 and the resulting COVID-19 disease severity remains to be fully understood.

Several studies have included asymptomatic and mild COVID-19 patients in cross-sectional analyses. For example, antibody responses and several cytokines/chemokines have been analyzed in asymptomatic versus symptomatic subjects^11^. Also, SARS-CoV-2-specific and functional memory T cells have been detected in recovered asymptomatic and mild COVID-19 patients^12^ or in recovered patients with undefined disease severity^13^. Such cross-sectional studies were critical to identify dysregulated immune factors that contribute to severe COVID-19. However, the isolated analysis of specific cellular immune subsets or of cytokines and antibody responses alone will only allow for a partial understanding of the coordination of the early immune response and trajectories following natural infection with SARS-CoV-2. Furthermore, due to different kinetics of immune responses among various patient groups, only a head-to-head comparison in a longitudinal, prospective study design can guarantee the comparability of observations between different study groups. This was only partially addressed in a recent longitudinal COVID-19 project, where the dynamic responses of various immune cells were investigated in asymptomatic, mild and hospitalized COVID-19 patients^7^. A comprehensive picture still remains incomplete because relevant observations and conclusions were based on various aggregated cohorts that were sampled at different time points, thus suffering from heterogeneous disease severity classification criteria, which were not strictly aligned.

In our longitudinal cohort characterized by a parallel and prospective study design, we sought to address the open questions regarding the kinetics, the differentiation, the quality and the evolution of early immune responses in mild versus asymptomatic and hospitalized COVID-19 patients following recent infection with SARS-CoV-2. To this end, we simultaneously analyzed 484 immune subsets and combinations of 36 lineage and functional markers in three flow cytometry panels, 24 serological cytokine/chemokine markers, serological antibody titers to SARS-CoV-2 Spike (S), S-receptor binding domain (RBD), S-N-terminal domain (NTD) and Nucleocapsid (N), angiotensin-converting enzyme 2 (ACE2) binding inhibition to S and S-RBD as surrogate for antibody neutralization capacity, and T-cell receptor beta (TCRb) repertoire sequencing in a longitudinal analysis with 220 samples. Key findings of our integrated analysis of all immune parameters suggested a highly-coordinated early-stage immune response including both innate and adaptive immune cells in mild non-hospitalized COVID-19 patients only. These early cellular response profiles were strongly correlated with a sufficient protective antibody production three weeks later. Both CD4^+^ and CD8^+^ T cell responses were mounted already very early after SARS-CoV-2 infection and showed an unexpected predominance of CD4^+^ SARS-CoV-2-specific TCRb clonotypes. Such a comprehensive, simultaneous and integrated analysis of various immune features in a longitudinal cohort using a systems-immunology strategy as we and others suggested^14,15^ helps to draw an unprecedented full picture of immune responses among mild non-hospitalized COVID-19 patients.

## Results

### Serological and whole blood count analysis distinguishing hospitalized from mild and asymptomatic COVID-19 patients

We established the longitudinal Predi-COVID cohort^16^ in Luxembourg during the first wave of the current pandemic with the aim to gain a deep and systematic understanding of the early antiviral immune response across the full spectrum of COVID-19 disease phenotypes (**Fig. 1a**). All patients received a diagnosis of COVID-19 confirmed by the SARS-CoV-2 PCR test through the national healthcare system and were then included into Predi-COVID with a delay of maximum 3 days after clinical PCR diagnosis. According to available clinical metadata, patients were stratified into asymptomatic (n=14), mild to moderate (n=63; referred to as “mild” patient group in **Fig. 1a** and thereafter), and hospitalized (n=15) subgroups for further analyses. All patients (n=92) were sampled on the day of inclusion (defined as “day 1”) and three weeks after inclusion (“day 21”). The group of mild COVID-19 patients also contained 11 patients with self-reported shortness-of-breath symptoms that could not be confirmed by a physician and therefore could not be classified as moderate patients following the NIH guideline. We also included control individuals (n=26) from patients’ households, who were sampled on day 1 and day 14. While the age range was not different for asymptomatic and mild patients as compared to household controls (**Supplementary Table 1, Fig. 1b**), hospitalized patients were older than mild patients (median, ∼57 vs. ∼38 years of age) and controls (**Fig. 1b**). The BMI was not different among any of the analyzed groups (**Supplementary Table 1**). In general, more males were included in each of the patient subgroups (between 57% and 69%), while only around 30% of household controls were male participants (**Supplementary Table 1)**. No comorbidity information was available for hospitalized patients and household controls. For the other patient groups, the prevalence of comorbidities (asthma, chronic hematologic disease, obesity and uncomplicated diabetes) was higher among mild than among asymptomatic patients (5% to ∼8% among mild vs. none among asymptomatic patients) (**Supplementary Table 1)**.

**Figure 1.**
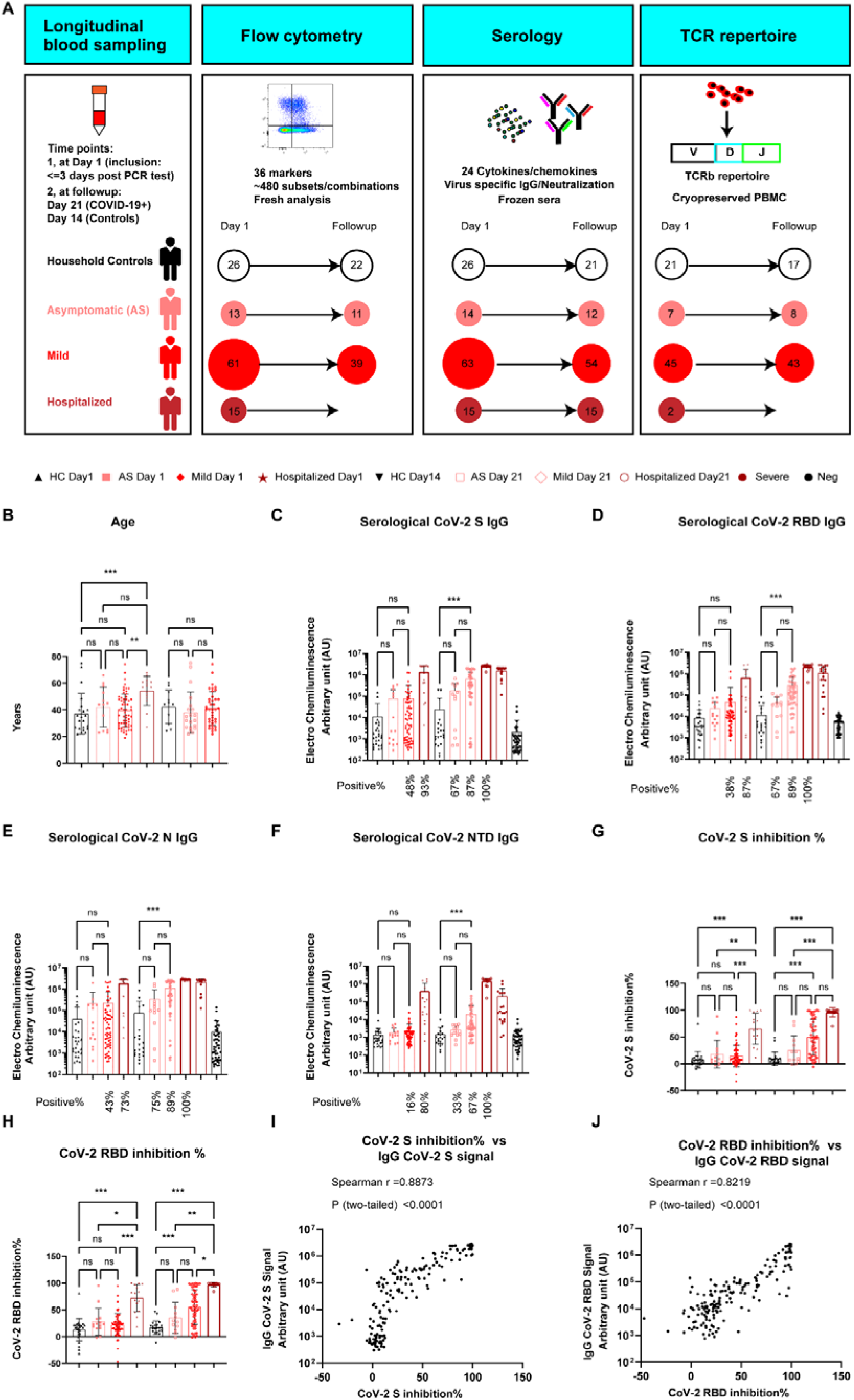
Cohort description and SARS-CoV-2 serological analysis of different groups. **A**, Sample numbers, longitudinal sampling scheme and experimental overview of different COVID-19 patient subgroups and household controls. AS, asymptomatic; HC, household controls. **B**, Scatter dot plots of age from each individual of different participant groups. **C, D, E, F**, SARS-CoV-2 spike-specific (**C**) or RBD-specific (**D**) or N-specific (**E**) or NTD-specific (**F**) IgG titers at day 1 inclusion for each group, or day 21 post inclusion of different patient subgroups or day 14 post inclusion of the household control group. The statistical test between non-hospitalized groups was based on the signals (AU). Only the positive percentages, rather than the signals of hospitalized samples were directly compared with that of other groups because different lots of abs were used (for the positive threshold calculation, refer to **Methods**). Neg, negative calibration sera from 2019 before the pandemic; Severe, positive sera from severe patients of another cohort of the local hospital (see **Methods**). **G, H**, Percentage inhibition by anti-SARS-CoV-2 Spike (**G**) or RBD (**H**) antibodies using a MSD pseudo-neutralization assay. The sera were diluted 50x before the measurement. **I, J**, Correlation between antibody titers and percentage inhibition of S (**I**) or RBD (**J**) antigens in all the COVID-19 patient samples. Spearman correlation was used for the analysis. Data represent individual values; Mean± standard deviation (S.D.); P-value in Figure **B-H** was determined by the Kruskal-Wallis (nonparametric) test and corrected using the Dunn’s multiple comparisons test. ns, not significant, *p<=0.05, **p<=0.01 and ***p<=0.001.

A whole blood count analysis was used to further characterize COVID-19 patients on day 1. We found no significant difference between asymptomatic and mild patients in any of the tested 17 general blood count parameters (**Supplementary Table 1, Supplementary Fig. 1**). However, hospitalized patients showed a remarkable difference compared to mild or asymptomatic patients as demonstrated in the principal component analysis (PCA) plot based on the analysis of whole-blood-count parameters (**Supplementary Fig. 1a**). In line with the reports by others^17^, the frequency of lymphocytes was substantially decreased in hospitalized patients compared to the two other patient groups (**Supplementary Fig. 1b**), while CRP was significantly elevated in hospitalized patients only (**Supplementary Fig. 1c**). Furthermore, hospitalized patients showed an increased number of white blood cells (WBC) (**Supplementary Fig. 1d**). This shift was mainly reflected by an increase in the number and frequency of both monocytes and granulocytes (**Supplementary Fig. 1e-h**). Both the red blood cell count and the hematocrit were modestly but significantly decreased, accompanied by a slightly higher number of platelets in hospitalized patients (**Supplementary Fig. 1i-j**). Although significantly enhanced, the number of platelets was still within the normal range in most of the hospitalized patients (**Supplementary Fig. 1j**).

As expected, on day 1 we did not observe a significant increase in IgG antibody levels to SARS CoV-2 S, CoV-2 RBD, CoV-2 NTD and CoV-2 N in the groups of asymptomatic and mild patients compared to controls **(Fig. 1c-f)**. In contrast, while only 48% and 43% of the mild patients showed slightly increased IgG levels, 93% and 75% of the hospitalized patients already displayed significantly enhanced IgG titers to CoV-2 S and N antigens respectively. Three weeks later, we observed a significant increase in IgG levels to all four antigens also in the mild patient group compared to household controls together with a further enhancement of IgG antibody levels in hospitalized patients. The positivity rate for IgG antibodies to CoV-2 S, CoV-2 RBD, CoV-2 NTD and CoV-2 N reached up to 89% among mild and 100% among hospitalized patients at day 21. Asymptomatic patients had a lower positivity rate for IgG against CoV-2 S/S-RBD (67%) and CoV-2 N (75%) than mild patients at day 21 (**Fig. 1c-e**). IgG antibodies against CoV-2 NTD were in general much lower in asymptomatic and mild patients, only reaching very high levels of 100% positivity in the hospitalized group at day 21 (**Fig. 1f**). Next, we tested the functional capacity of the induced antibodies against SARS-CoV-2 in a surrogate virus neutralization assay. The assay analyzes the capacity of antibodies to inhibit the binding of labelled recombinant ACE2, the human receptor for SARS-CoV-2, to CoV-2 S or CoV-2 RBD in a multiplex high-throughput format. In line with the serology findings on day 1 (**Fig. 1c, d),** hospitalized patients already showed blocking antibodies that interfered with ACE2 binding to CoV-2 S or CoV-2 RBD at this early stage **(Fig. 1g, h)**. On day 21, also mild patients had developed a significant inhibitory serologic capacity to block ACE2 binding to CoV-2 S or CoV-2 RBD relative to household controls. Such an increase in ACE2 blocking was not seen in the asymptomatic group **(Fig. 1g, h)**, which was also lower in IgG titers against CoV-2 S or CoV-2 RBD (**Fig. 1c, d)**. In line with this notion and as reported by others^18^, IgG antibody titers against both CoV-2 S and CoV-2 RBD, were highly correlated (spearman r=0.89 and 0.82 for CoV-2 S and RBD, respectively) with the inhibitory capacity of sera across all patient categories of COVID-19 severity (**Fig. 1i, j**).

### Early-stage highly-coordinated innate and adaptive immune responses in mild COVID-19

As shown above, clinical and routine laboratory data as well as in depth serologic profiling of SARS-CoV-2-specific antibody responses already distinguished hospitalized COVID-19 patients from mild or asymptomatic patients and control individuals. However, these profiling analyses are not sufficient to further differentiate non-hospitalized clinical phenotypes of COVID-19. Thus, we aimed to explore the full complexity of innate and adaptive cellular immune signatures that orchestrate the early response to SARS-CoV-2 infection across the full spectrum of COVID-19 disease phenotypes. We systematically investigated 484 cellular immune subsets or combinations of various lineage and functional markers by three different staining panels using 18-color flow cytometry (for general gating strategy, see **Supplementary Fig. 2**; for cellular markers analyzed, refer to **Supplementary Table 2**) on day 1 and at the follow-up visit (day 14 for controls or day 21 for patients). When compared with household controls, asymptomatic patients displayed no obvious change in all the analyzed 484 immune profiles at day 1 (**Supplementary Fig. 3a**). Interestingly, at day 21, ICOS^+^ CD8 T cells were the only significantly changed immune subset with a decrease in the frequency among CD8 T cells from asymptomatic patients (**Supplementary Fig. 3b, c**). Although primarily known for its critical role in CD4 T cell and lymph node germinal center formation, studies in ICOS-deficient patients have also indicated a role for ICOS in CD8 effector functions during primary antiviral immunity^19,20^.

We next used PCA to show that deep immune profiling was only able to partition hospitalized patients at day 1 from all other groups investigated, but not mild COVID-19 patients from any household control (day 1 and day 14) (**Fig. 2a**). Then we asked whether specific immune subsets were differentially present in mild COVID-19 patients compared to age-matched household controls at day 1 (**Fig. 2b**). We observed differences in the frequency of several CD8 T cells subsets, such as Ki67^+^, CD38^+^, and HLADR^+^CD38^+^, representing proliferating, activated and antigen-specific responsive CD8 T cells respectively that were significantly enhanced in mild COVID-19 patients (**Fig. 2c-e**). The profile included an increase of both Tbet-dependent (Tbet^+^Ki67^+^) and -independent (Eomes^+^Ki67^+^) responsive CD8 T cells (**Fig. 2f-h**). Also the fraction of proliferating CD4 T cells, especially, Th1-responsive (Tbet^+^Ki67^+^) CD4 T cells was already enhanced early on in mild patients on day 1 (**Fig. 2i, j, Supplementary Fig. 4a**). In parallel, the frequency of antigen presenting cells (APCs) and antibody secreting cells, such as mature dendritic cells (HLADR^+^CD38^high^DCs) and short-lived plasmablasts (CD27^+^CD38^high^), was increased in mild patients (**Fig. 2k, l, m, Supplementary Fig. 4b, 4c**). Notably, using the unique power of our longitudinal cohort design and the simultaneous comprehensive analysis of both cellular immune subsets and serological responses, allowed the successful prediction of anti-SARS-CoV-2 antibody responses at day 21. The frequency of activated CD38^+^ CD8 T cells and of mature DCs measured at day 1 was highly predictive for the degree of the serological titers of anti-SARS-CoV-2 N IgG at day 21 among asymptomatic and mild patients (**Fig. 2n, o**). On the contrary, it is noteworthy that neither the frequency of CD38^+^ cells among CD8 T cells nor of mature DCs at day 1 was significantly correlated to the even higher titers of SARS-CoV-2 N IgG (**Fig. 2l, m**) in the group of hospitalized COVID-19 patients at day 21 (**Fig. 2p, q**). This finding indicates that the progression and deterioration of COVID-19 is averted only in the presence of a highly coordinated interplay of early cellular innate and adaptive immune responses, which are strongly correlated to the subsequent production of protective antibody titers against SARS-CoV-2.

**Figure 2.**
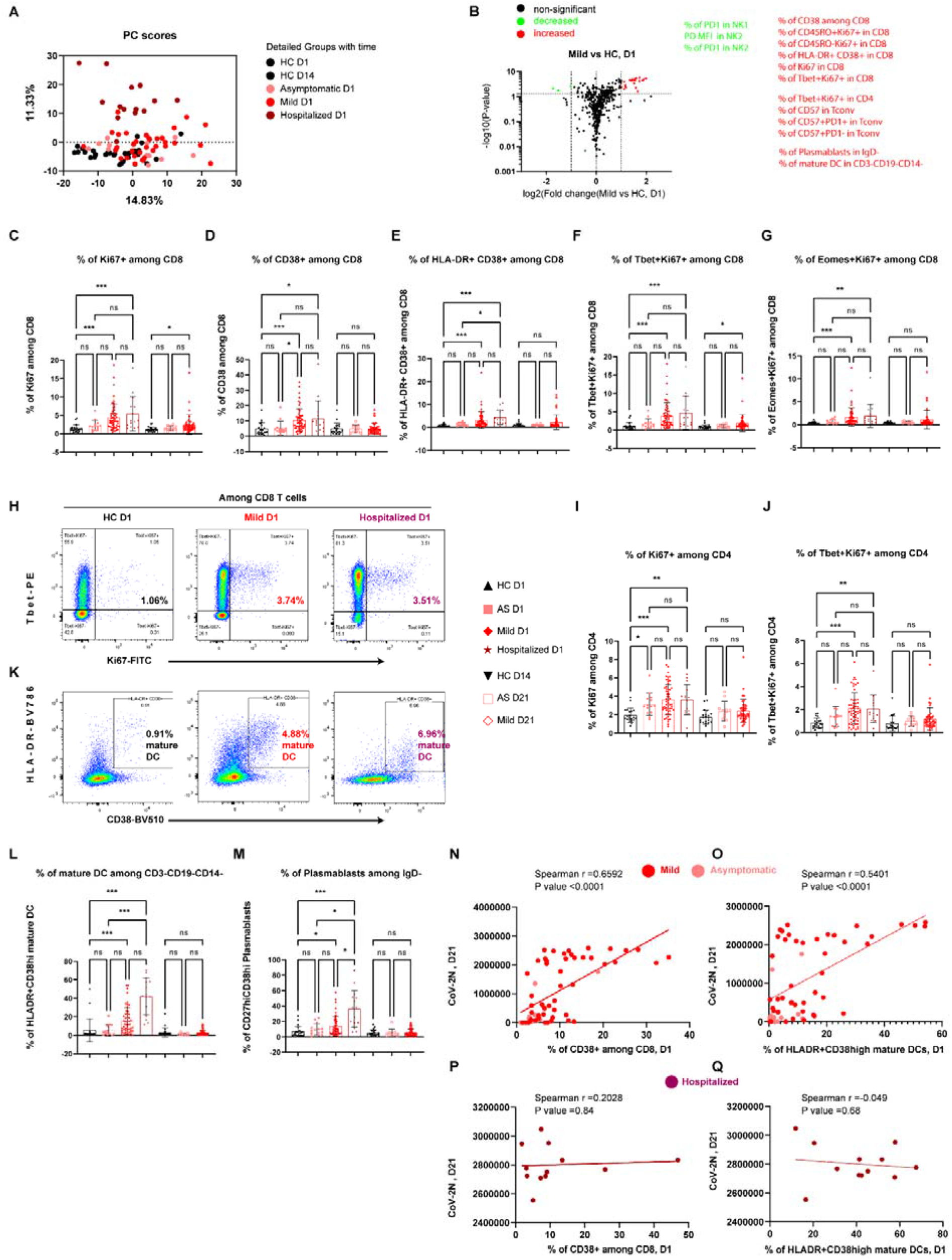
Early-stage coordinated responses of CD4, CD8, mature DC and Plasmablasts in mild COVID-19 patients. **A**, PCA plots of the samples from different patient groups at day 1 of inclusion and the house hold controls at day 1 and day 14 post inclusion. The analysis was based on 484 immunological features/subsets analyzed by multi-panel and multi-color flow cytometry. **B**, Volcano plots of different immune features in patients with mild symptoms vs. household controls at day1 of inclusion. The selected list of significantly increased or decreased subsets (p<=0.05 and change fold >=2) were marked in red or green, respectively. Tconv, FOXP3^-^CD4 conventional T cells. **C, D, E, F, G**, Frequency of Ki67^+^ (**C**), CD38^+^ (**D**), HLA-DR^+^CD38^+^ (**E**), Tbet^+^Ki67^+^ (**F**), EOMES^+^Ki67^+^ (**G**) among CD8 T cells from different groups at different time points. AS, asymptomatic; HC, household controls; D1/D14/D21, day 1/day 14/day 21. **H, K**, Representative flow cytometry plots of the expression of Tbet and Ki67 (**H**) or the expression of CD38 and HLA-DR (**K**) on CD8 T cells from either household controls, mild or hospitalized patients at day 1 of inclusion. **I, J**, Frequency of Ki67^+^ (**I**) and Tbet^+^Ki67^+^ (**J**) among CD4 T cells. **L**, Frequency of HLA-DR^+^CD38^high^ mature Dendritic cells (DC) among CD3^-^CD19^-^CD14^-^ cells. **M**, Frequency of CD27^high^CD38^high^ plasmablasts among CD3^-^CD19^+^IgD^-^ B cells. **N, P**, Correlation between the frequency of CD38^+^ among CD8 at day 1 and anti-SARS-CoV-2N-specific IgG titers at day 21 following inclusion from asymptomatic and mild patients (**N**) or from hospitalized patients (**P**). Spearman correlation was used for the analysis. **O, Q**, Correlation between the frequency of mature dendritic cells among CD3^-^CD19^-^CD14^-^ cells at day 1 and anti-SARS-CoV-2N-specific IgG titers at day 21 following inclusion from asymptomatic and mild patients (**O**) or from hospitalized patients (**Q**). Data represent individual values; Mean± standard deviation (S.D.); P-value in Fig. **C-G, I-M** was determined by the Kruskal-Wallis (nonparametric) test and corrected using the Dunn’s multiple comparisons test. ns or unlabeled, not significant, *p<=0.05, **p<=0.01 and ***p<=0.001.

Now switching to day 21 after inclusion, we followed the evolution of the early cellular immune response. On day 21, mild COVID-19 patients were characterized by an enhanced cytotoxic CD8 T cell (GZMB^+^) response, especially of terminally differentiated responsive CD8 T cells (CD45RO^-^ Ki67^+^) (**Supplementary Fig. 5a-c**). CD4 T cells also showed similar changes after three weeks. The frequencies of CD4 T cells expressing CD57 and GZMB as well as of CD45RO and CD57 double-positive CD4 T cells were also significantly enhanced in mild patients compared to household controls (**Supplementary Fig. 5d-f**). Notably, the frequency of GZMB^+^ CD4 cytotoxic T cells showed a trend to be elevated (p=0.053, Kruskal-Wallis test including multiple-group correction) already on day 1 (**Supplementary Fig. 5e**). The CD57 expressing CD4 T cells detected on day 21 appeared to be mainly cytotoxic effector cells since the percentage of GZMB^+^CD57^+^ cells was also significantly enhanced among CD4 T cells (**Supplementary Fig. 5g**). Although CD57 is regarded and known as a T-cell senescence marker, CD57^+^ T cells, similar to PD-1^+^ T cells^21^, were apparently still functional during the acute phase of COVID-19, thus likely contributing to a sufficient control of the infection in the mild patient group.

### Early-stage impaired innate immunity in hospitalized, but not mild patients

In the analyses presented hitherto, we parsed primarily early immune signatures in mild COVID-19 patients in relation to household controls on day 1 and day 21. Yet, the analysis of early cellular responses and later antibody responses showed a positive correlation only in mild and asymptomatic, but not in hospitalized patients (**Fig. 2n-q**). This finding prompted us to further analyze this aspect. Thus, we asked whether any additional early immune signatures observed in mild patients were significantly different from hospitalized patients and determined the immune signatures that were significantly upregulated or downregulated in mild versus hospitalized COVID-19 patients on day 1 (**Fig. 3a**). As shown in the volcano plot analyses, major differences were present primarily among innate immune cells, such as monocytes, dendritic cells (DC) and natural killer (NK) cells, and to a lesser extent also among B and T cells. Compared with hospitalized patients, mild COVID-19 patients showed a much higher frequency (∼40% in mild patients vs. ∼10% in hospitalized patients) of non-classical monocytes (ncMono, HLADR^+^CD38^-^)^22^. The non-classical monocytes were not only higher in frequency among the mild patients, but expressed also higher levels of critical functional markers, such as CD86/CD80 double-positivity (**Fig 3b-d**), PD-L1 and CD13 (**Supplementary Fig. 4d, Supplementary Fig. 6a, b**). Similar to monocytes, the frequency of antigen-presenting cells (APC) such as plasmacytoid DC (pDC) and myeloid DC (mDC) was significantly higher in mild patients versus hospitalized ones (**Fig. 3a, d-f**). It is noteworthy that the frequency of ncMono and mDC (**Fig. 3b, f**) was also slightly lower in mild patients versus household controls, indicating a disease severity-related effect and further supporting the involvement of both cell types in early protective immune responses of COVID-19. Although mature DC were higher in both mild and hospitalized patients (**Fig. 2l**), the frequency of CD86^-^CD80^+^ functional cells (**Fig. 3g, h**) and of CD13^+^ cells (**Supplementary Fig. 6c**) among total DC was decreased in hospitalized patients only, thus indicating a reduction in phagocytic and antigen-presenting capacity of individual DC^23^. These findings signify one of the advantages of our study, where we analyzed not only lineage, but also functional markers, thus allowing a better comprehension and interpretation of seemingly conflicting results. In line with the notion of reduced APC functions, the downstream events of APC activating responses, the frequency of activated CD4 T cells (CD27^+^ICOS^+^) and the ICOS MFI among CD8 T cells were decreased only in hospitalized, but not mild patients (**Supplementary Fig. 6d, e**). Furthermore, the frequency of NK cells was also significantly decreased only in hospitalized, but not in mild patients relative to household controls (**Fig 3i, j**). In line with the overall compromised innate immune cell profile, critical senescence and exhaustion markers such as KLRG1 and PD-1 were enhanced among several subsets of NK cells in hospitalized patients only (**Fig. 3k, Supplementary Fig. 4e, Supplementary Fig. 6f, g**).

**Figure 3.**
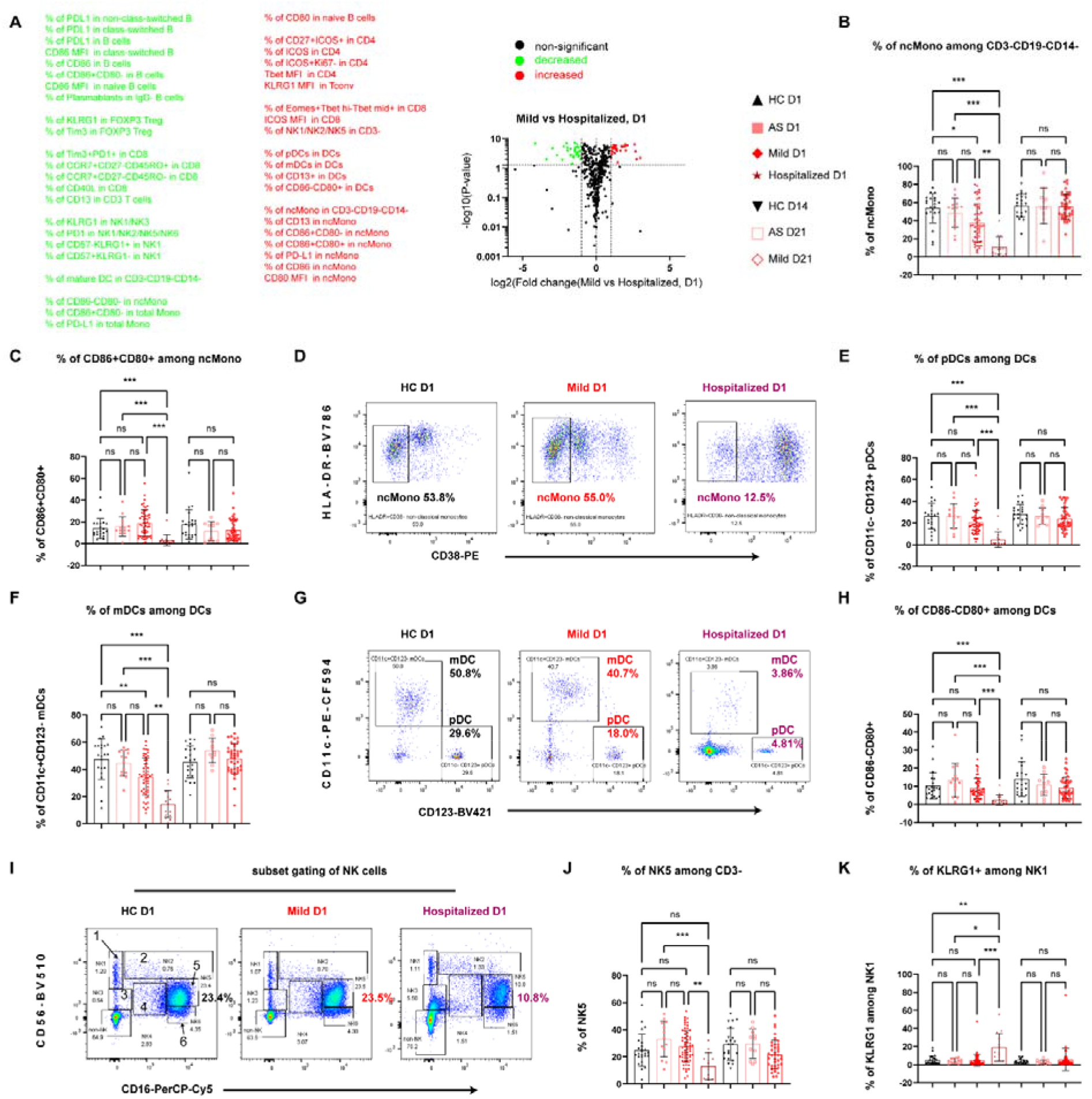
Impaired early-stage responses of non-classical monocytes, DC and NK cells distinguishing hospitalized COVID-19 patients from mild patients. **A**, Volcano plot showing the comparison of the frequency of different immune subsets in mild versus hospitalized patients at day 1. The selected list of significant increased or decreased subsets (p<=0.05 and change fold >=2) were marked in red or green, respectively. AS, asymptomatic; HC, household controls; D1/D14/D21, day 1/day 14/day 21. **B**, Proportions of HLA-DR^+^CD38^-^ non-classical monocytes (ncMono) among CD3^-^CD19^-^CD14^-^ cells. **C, E, F**, Frequency of cells expressing CD86^+^CD80^+^ among ncMono (**C**) or pDC (**E**) or mDC (**F**) among total DCs. **D, G**, Representative flow-cytometry plots of the expression of HLA-DR and CD38 (**D**) or the expression of CD11c and CD123 (**G**) from different groups. **H**, Frequency of CD86^-^CD80^+^ cells among DCs. **J, K**, Frequency of NK5 among CD3^-^ cells (**J**) or of KLRG1 among NK1 (**K**). **I**, Representative flow-cytometry plots of the expression of CD56 and CD16. Gating strategy to define six subsets of NK cells. Enlarged number from 1 to 6 represents various NK subsets. Data represent individual values; Mean± standard deviation (S.D.); P-value was determined by the Kruskal-Wallis (nonparametric) test and corrected using the Dunn’s multiple comparisons test. ns or unlabeled, not significant, *p<=0.05, **p<=0.01 and ***p<=0.001.

In contrast to the compromised innate immune cell compartment, the expression levels of CD86 and of PD-L1 among class-switched memory B cells were substantially enhanced in hospitalized COVID-19 patients versus both household controls and mild patients at day 1 (**Supplementary Fig. 6h,i**). Considering these results together with the high SARS-CoV-2-specific IgG levels, the ACE2 blocking capacity of patient serum and the high frequency of plasmablasts, we concluded that antibody-secreting cells were not impaired in both mild and hospitalized patients at the very early stage (on day 1), thus leading to a robust antibody response in both patient groups after three weeks.

We next sought to understand whether the impaired innate immune cell response in hospitalized patients was paralleled by early deviated CD8 T cell profiles. Although the intensity (MFI) of ICOS on CD8 T cells was decreased (**Supplementary Fig. 6e**), the frequency of ICOS^+^ CD8 T cells was unchanged (**Supplementary Fig. 3c**) and the frequency of CD40L^+^ and PD-1^+^GZMB^+^ cells among CD8 T cells was even significantly enhanced in hospitalized patients on day 1 (**Supplementary Fig. 6j, k**). Furthermore, since the frequency of CD8 T cells expressing other key functional markers, such as Ki67 and CD38, as well as the frequency of CD8 T cells co-expressing HLA-DR and CD38, Tbet and Ki67, as well as Eomes and Ki67 was not decreased in hospitalized patients (**Fig. 2c-g**), the functional antiviral capacity of CD8 T cells was most likely equally robust in hospitalized and mild COVID-19 patients. Overall, these data indicate that the major deficiencies observed in hospitalized COVID-19 patients on day 1 were impaired innate immune cells and APC functions rather than adaptive T- and B-cell functions.

The differences in cellular immune signatures described above between mild and hospitalized patients were only significant on day 1, but not after three weeks on day 21. This indicates that various immunological signatures, which are reduced in hospitalized patients on day 1, are unique to the mild clinical phenotype of COVID-19 and may thus be crucial protective factors at an early stage of the disease.

### Early-stage temporary and reversible elevation of IP10 and IFNb in mild COVID-19 patients

To gain further insight into the coordinated early immune response of COVID-19, we analyzed 24 different cytokines, chemokines and growth factors on both day 1 and day 21 in sera of all patient and control groups. Interestingly, at day 1, we observed increased levels of interferon gamma-inducible protein 10 (IP10/CXCL10) in hospitalized patients and in the mild patient group (**Fig. 4a, b**). Unexpectedly, a similar regulation was found for the type I-interferon IFNb, which was previously reported to be undetectable in severe COVID-19 patients at around 10 days after symptom onset^24^. Both hospitalized and mild patients showed a significant increase of IFNb compared to controls at day 1 (**Fig. 4c**). While the levels of IP10 and IFNb showed only a temporary and reversible increase among mild patients, declining to normal levels at day 21, this was not the case in hospitalized patients, where both IP10 and IFNb levels remained elevated after three weeks (**Fig. 4c**). These results point to a crucial and dynamic role of IP10 and IFNb, which is tightly regulated during the early stage of protective immune responses in COVID-19 patients. This notion is also supported by the fact that levels of IP10 and IFNb were significantly correlated with the frequency of mature DCs among all the analyzed patients at day 1 of the Predi-COVID study (**Fig. 4d, e**). On day 21, none of the 24 circulating immune analytes showed a significant change in the mild patient group versus household controls at day 14 (**Supplementary Fig. 7a**). We also could not observe any significant change in the tested cytokine/chemokine levels among asymptomatic patients, neither on day 1 nor on day 21 (**Supplementary Fig. 7b, c**). The increase of IP10 in mild and hospitalized COVID-19 patients appears to be independent of IFNg, which was only elevated in the hospitalized patient group (**Fig. 4c, f, g**) and remained significantly higher in hospitalized patients, but still within the normal range for most of the patients (**Fig. 4g**). Overall, the IP10 and IFNb signature in mild COVID-19 patients was characterized by early dynamic changes with a strong increase at day 1 and a contraction to normal levels at day 21, while the levels in hospitalized patients remained high.

**Figure 4.**
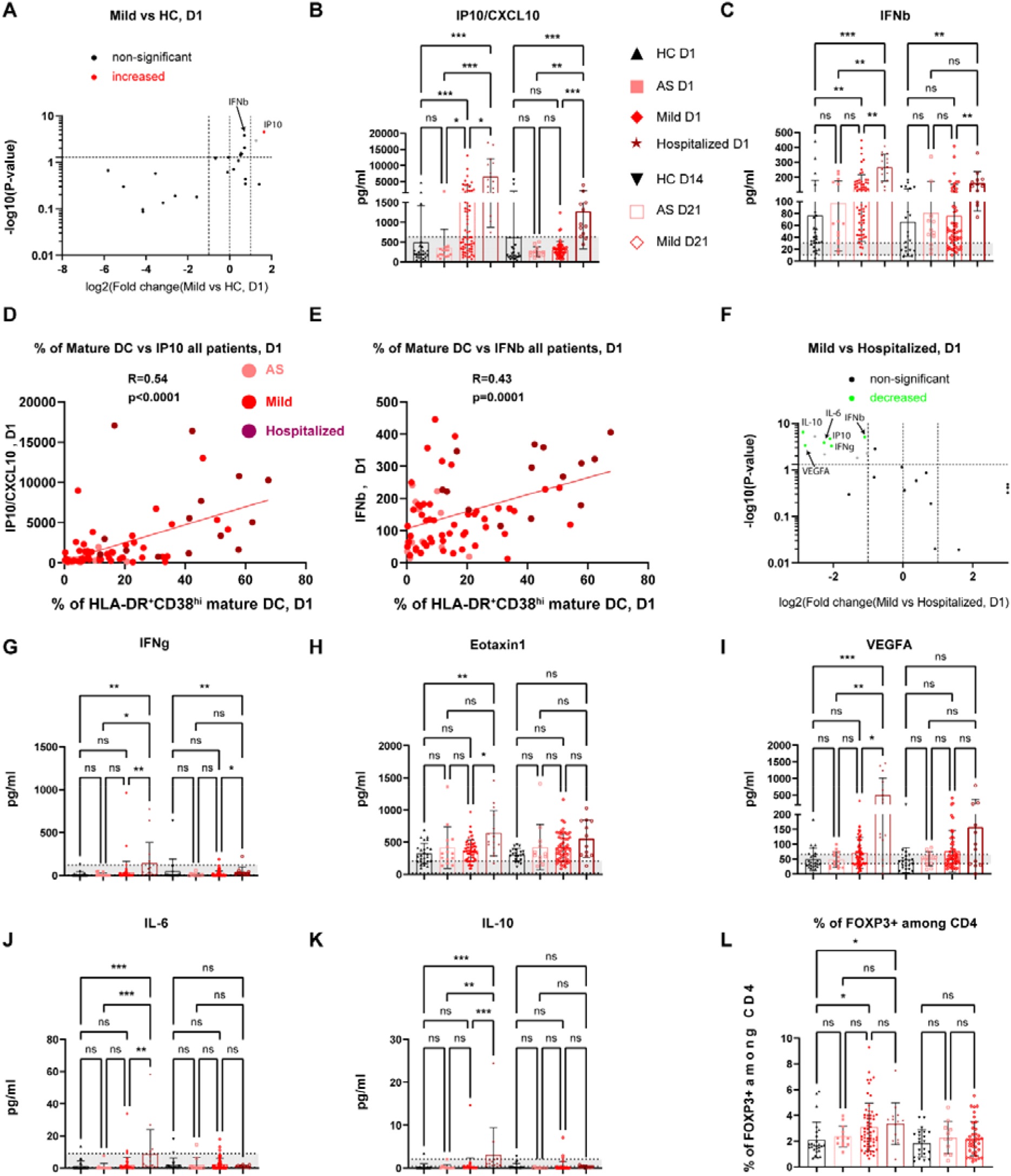
Early-stage transient cytokine responses in mild COVID-19 patients. **A**, Volcano plot showing the serological cytokine/chemokine responses in mild patients relative to household controls (HC) at day 1 of inclusion. Significantly increased or decreased cytokines/chemokines/growth factors (p<=0.05 and change fold >=2) were marked in red or green, respectively. The gray dot represents the analytes showing a significant change but displaying values lower than the reported normal physiological levels even in control groups. **B, C**, Scatter dot plots of serological levels of IP10 (**B**) and INFb (**C**) of different groups at day 1 or day 14 or day 21 following inclusion. AS, asymptomatic; HC, household controls; D1/D14/D21, day 1/day 14/day 21. **D, E**, Correlation between the frequency of mature DC and IP10 (**D**) or IFNb (**E**) at day 1 of inclusion. R, Pearson correlation coefficient. Different groups were marked by different indicated colours. **F**, Volcano plot showing the responses of serological cytokines/chemokines of mild patients versus hospitalized patients at day 1 of inclusion. **G, H, I, J, K**, Scatter dot plots of IFNg (**G**), Eotaxin1 (**H**), VEGF A (**I**), IL-6 (**J**) and IL-10 (**K**) of different participant groups. **L**, Frequency of FOXP3^+^ Treg cells among CD4 T cells. Data represent individual values; Mean± standard deviation (S.D.); P-value was determined by the Kruskal-Wallis (nonparametric) test and corrected using the Dunn’s multiple comparisons test. ns or unlabeled, not significant, *p<=0.05, **p<=0.01 and ***p<=0.001. Gray shading indicates the reported normal range for different cytokines/chemokines.

With regard to other circulating immune factors, we found a significant increase in plasma levels of eosinophil chemotactic protein (eotaxin-1/CCL11), vascular endothelial growth factor A (VEGFA), IL-6 and IL-10 only in hospitalized patients at day 1 (**Fig. 4h-k**). The enhanced levels of IL6 and the regulatory cytokine IL10 in hospitalized versus mild patients on day 1 were still mostly seen within the normal range (**Fig. 4j, k**). The increased level of IL10 was in line with an increased percentage of FOXP3^+^ Tregs in both mild and hospitalized patients at day 1 (**Fig. 4l**). In conclusion, on day 21 after inclusion into the study, most of the analyzed cytokines/chemokines that were elevated early in mild patients had returned to normal levels and waned in hospitalized patients. Only IFNg, IFNb and IP10/CXCL10 remained higher in hospitalized patients than mild patients and controls on day 21 (**Supplementary Fig. 7d**). The only exception was the Th2 cytokine IL5, which was not different on day 1, but modestly increased in mild patients at day 21, while hospitalized patients exhibited very low IL5 levels at this point (**Supplementary Fig. 7e**).

### Early-stage dominant expansion of CD4+ SARS-CoV-2-specific T cells in mild patients

To identify the T-cell response on a broader scale, we performed TCRb sequencing analysis among 45 mild patients versus 8 asymptomatic subjects and 21 household controls on days 1 and 21. Aging has a strong impact on the TCR repertoire^25^. As expected, sample clonality, the inverted normalized diversity index, was significantly correlated with the age of all analyzed subjects (**Fig. 5a**). Since a decrease in TCR diversity was previously associated with aging and impaired immunity against influenza virus infection and other diseases^26,27^, we sought to compare the TCR diversity between different groups. The productive clonality of the sequenced TCRb repertoire was increased in mild patients at day 21 versus household controls (**Fig. 5b**). At day 21, only the usage of one specific V gene (TCRBV06-07) was significantly underrepresented in mild patients compared to household controls (**Fig. 5c**). Notably, the SARS-CoV-2-specific T-cell clonotypes were substantially expanded (∼6 times higher than in household controls) among mild patients already at day 1, as reflected by clonal breadth and depth ^28^, and maintained at day 21 (**Fig. 5d, e**). These results indicate a key functional role of early-responsive SARS-CoV-2-specific T cells in mild COVID-19 patients. Completely unexpected, mainly CD4 SARS-CoV-2-specific T cells were expanded among mild patients at day 1, with an average frequency of CD4 SARS-CoV-2-specific TCR clonotypes that was six times higher than that of CD8 T cells among mild patients (**Fig. 5f-h**). This finding was in line with a trend for increased frequency of GZMB^+^ cells among CD4 Tconv cells, but not among CD8 T cells in mild patients versus controls on day 1 (**Supplementary Fig. 5a, e**). At day 21, CD4 SARS-CoV-2 specific TCR clonotypes continued to dominate over CD8 clonotypes to a similar extent in mild patients (**Fig. 5g, h**). The expansion of CD4 or CD8 SARS-CoV-2-specific T cells was highly correlated with the frequency of responsive ICOS^+^Ki67^+^ cells among total CD4 T cells or total CD8 T cells in both asymptomatic and mild patient groups at day 1 (**Fig. 5i, j**). These data highlight a crucial role of early-expanding SARS-CoV-2-specific T cells, especially of the CD4^+^ phenotype, for subsequent coordinated antiviral immune responses. Thus, our findings are not only in line with, but add significantly novel aspects to earlier studies that identified SARS-CoV-2-spectific T cells early after diagnosis, but without correlating their findings to a severity stratification of COVID-19 patients nor performing further sub-analysis of CD4 and CD8 T cells^29^.

**Figure 5.**
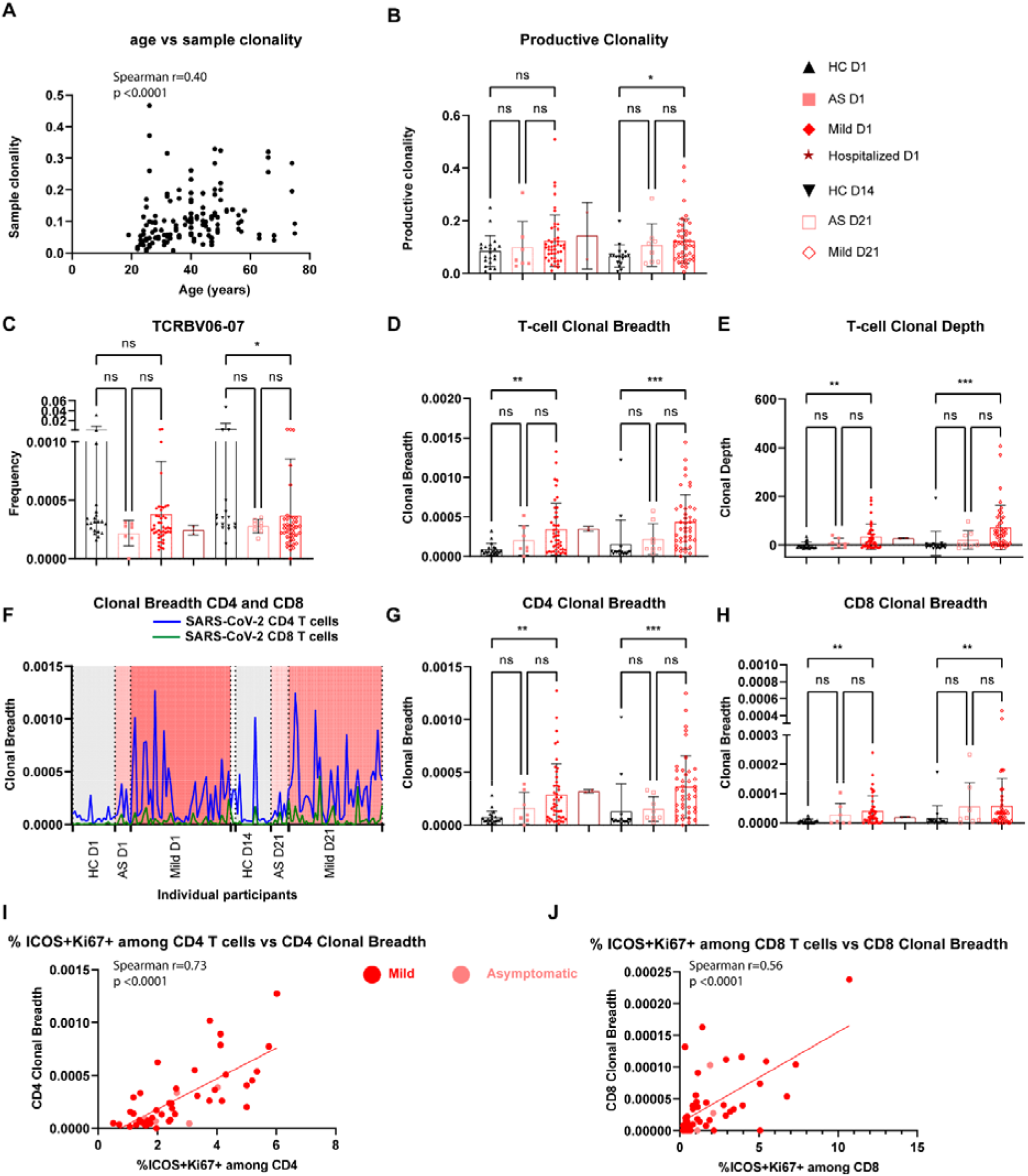
Early-stage expansion of SARS-CoV-2-specific TCR clonotypes in mild COVID-19 patients. **A**, Correlation between sample clonality and age of corresponding participants of the samples (n=144) from all the groups. r, Spearman correlation coefficient. **B**, Productive clonality (inverted normalized diversity index) of different groups. Different groups were marked by different indicated colours. AS, asymptomatic; HC, household controls; D1/D14/D21, day 1/day 14/day 21. **C**, The usage frequency of the TCRbeta V06-07 gene among different groups. **D, E**, Clonal breadth (**D**): Relative frequency of SARS-CoV-2-specific T cell clonotypes among unique productive rearrangements; Clonal Depth (**E**), expansion extent of SARS-CoV-2-specific T cell clonotypes. **F**, Clonal breadth of CD4 and CD8 SARS-CoV-2-specific TCR clonotypes among each individual participant from different groups. The values from every individual participant were linked through lines. The two D1 hospitalized patients were posited between Mild D1 and HC D14 (the gap between the dashed lines), but not labelled. **G, H**, Clonal breadth of CD4 (**G**) or CD8 (**H**) SARS-CoV-2-specific TCR clonotypes among different groups. **I, J**, Correlation between SARS-CoV-2-specific TCR clonal breadth and percentages of ICOS^+^Ki67^+^ among CD4 T cells (**I**) or among CD8 T cells (**J**) in asymptomatic and mild patients at day 1. Correlation coefficient was based on Spearman correlation. P-value was from the two-tailed test. Data represent individual values; Mean± standard deviation (S.D.); P-value from the Fig. **B-E, G-H** was determined by the Kruskal-Wallis (nonparametric) test and corrected using the Dunn’s multiple comparisons test. ns or unlabeled, not significant, *p<=0.05, **p<=0.01 and ***p<=0.001.

## Discussion

In May 2020, a mass PCR screening program was implemented on a population-wide level in Luxembourg^30^, which allowed us to get unique access to PCR-positive asymptomatic and mild non-hospitalized COVID-19 patients and to prospectively recruit them into the longitudinal Predi-COVID study that was initiated simultaneously^16^. This empowered us to analyze a comprehensive picture of distinct early-stage protective immune signatures in mild COVID-19 patients versus hospitalized patients, asymptomatic individuals and healthy controls during the first waves of the pandemic. We evaluated more than 500 cellular and soluble markers of the immune response in the peripheral blood of all study participants twice in the early time window after infection. Furthermore, we performed systematic TCRbeta variable chain sequencing at the identical time points in our cohort. Thus, our work provides an important resource based on the unique opportunity to fully explore and understand all essential facets of the early-stage and dynamic immunological changes following recent SARS-CoV-2 infection in mild COVID-19 patients, using an unbiased and prospective approach.

So far, the immune response in COVID-19 patients has only been investigated in a few longitudinal cohort studies^17,31–34^. These studies concentrated on different time windows and they usually put their major focus on one or two selected immunological aspects, which makes it challenging to directly compare them with our results. In line with another recent longitudinal study from the UK^7^, we also observed enhanced early-stage CD8 T cells and plasmablast responses in mild COVID-19 patients. In contrast to that published work, our current study now provides not only information on the number and frequency of a wide spectrum of immune subsets in peripheral blood, but also on the functionality of individual immune cell types. We found both early-stage Tbet-dependent and - independent CD8 T cell responses among mild COVID-19 patients. Distinct from the UK cohort^7^, we observed robust early-stage responses of CD4 T cells with a profoundly enhanced frequency of type-I-IFN-dependent Tbet^+^Ki67^+^ CD4 T cells. As additional unique result of our study, mature DC expressing all key functional markers were strongly enhanced early on in mild but not hospitalized patients. The notion that early coordinated DC and CD4/CD8 T cell responses have indeed a functional role in SARS-CoV-2-specific immunity was further supported by the correlation between mature DC and CD38^+^CD8 T cells at early stage (day 1) with antibody responses three weeks later (day 21). Consistent with the observed early-stage CD4 and CD8 T cell responses, we found a substantial expansion of SARS-CoV-2-specific T-cell clonotypes, predominantly of CD4 T cells, in mild COVID-19 patients already at day 1. Since we observed enhanced DC and coordinated CD4 and CD8 T cell responses very early on among mild COVID-19 patients, at most three days after PCR diagnosis, the concept of bystander CD8 T cell responses^7^ might need to be adapted. Our findings suggest that appropriate and highly coordinated early-stage DC and antigen-specific T cell responses are crucial for guiding the development of a protective adaptive immune response at later stage, which is key for a long-term favorable outcome in non-hospitalized mild COVID-19 patients.

Another important observation that is very much in line with the coordinated early-stage DC and antigen-specific CD4 and CD8 T cell responses was the strong early induction of the type-I interferon IFNb in both mild and hospitalized COVID-19 patients. However, in contrast to hospitalized patients, the early rise of IFNb levels in mild patients was followed by a decrease to normal levels three weeks later. Such a contraction of IFNb was not seen in hospitalized patients, where the levels remained high three weeks later, thus indicating a possible dysregulation in more severe COVID-19 phenotypes. Indeed, previous studies had demonstrated impaired type-I interferon responses in severe COVID-19 patients^24,35,36^. Our current results on early induction and contraction of IFNb levels in mild COVID-19 patients were further confirmed by a strong correlation between the frequency of mature DC, one of the main producers of type-I interferon^37^, and the circulating IFNb levels on day 1. Our data suggest a protective role of the type-I interferon IFNb that may be crucial during the very early stage of COVID-19 after recent exposure to SARS-CoV-2. The results of an early time-dependent induction of IFNb in our study are in line with studies on genetic and autoimmune defects in the type-I IFN pathway that were correlated to severe COVID-19^24,35,36^. These findings have prompted to propose early and transient intervention with recombinant type-I IFN such as IFNb as a treatment option in severe COVID-19 patients^38^. Parallel to IFNb, we observed substantially enhanced early IP10/CXCL10 levels without other signs of systemic inflammation in mild patients. After three weeks, IP10 levels had declined to the normal range in mild, but not in hospitalized COVID-19 patients, thus indicating that a temporary early-stage enhancement of IP10 may be beneficial in mild COVID-19. IP10, previously known to exclusively bind to CXCR3, has recently been identified as a high-affinity agonist for the anti-inflammatory atypical chemokine scavenger receptor ACKR2/D6^39,40^. Thus, during the coordinated early anti-SARS-CoV-2 immune response in mild patients, IP10 might play a role in limiting and resolving inflammatory responses. The longer-lasting high levels of IP10 are not unique to severe COVID-19, but occur also in other infectious diseases^41^, including SARS, where high levels of IP10 were maintained for at least two weeks^42^. Our observations are also in line with previously reported enhanced IP10 levels in symptomatic versus asymptomatic patients during the acute phase of COVID-19^11^ and sustained levels in hospitalized patients^43^.

Notably on day 1, hospitalized COVID-19 patients showed significantly increased levels of vascular endothelial growth factor A (VEGFA), known to be critical in acute lung injury^44^, and of eosinophil chemotactic protein (eotaxin-1/CCL11). Enhanced VEGFA mRNA expression has previously also been demonstrated in the bronchial alveolar lavage fluid of two COVID-19 patients^45^. Furthermore, a significant elevation of VEGFA and eotaxin-1 among both mild and severe COVID-19 patients relative to controls, although without significant difference between the two groups, was reported in a small scale pilot study^46^ and anti-VEGF medication has been investigated to treat COVID-19 in a phase-II clinical trial^47^. Our findings strongly suggest that VEGFA and eotaxin-1 are indeed differentially regulated during the early phase of the immune response in hospitalized, i.e., more severe COVID-19 patients.

Another crucial observation of our study was an early-stage signature of reduced frequency and functional impairment of innate immune cells, such as non-classical monocytes (ncMono), DC (pDC and mDC) and NK cells, in hospitalized COVID-19 patients only. Both the frequency of ncMono and the expression of their key functional markers, such as CD80, CD86, PD-L1 and CD13, were significantly reduced in hospitalized patients. Notably, although the frequency of mature DC was induced early on in both mild and hospitalized patients, their functional subsets, such as CD86-CD80+ cells among total DC, were substantially reduced only in the hospitalized patients. In addition, similar findings were also made in hospitalized patients regarding the reduced frequency of several NK cell subsets, paralleled by a substantial enhancement in the expression of the inhibitory and terminal differentiation marker KLRG1 on NK cells. Thus, our data of impaired innate immune cell signatures in hospitalized, but not mild COVID-19 patients confirm previous findings of impaired innate immunity in severe or critically ill COVID-19 patients^48,49^. Furthermore, our results are in line with the decreased frequency of pDC in hospitalized patients versus control subjects, which was revealed through a longitudinal single-cell RNA sequencing analysis^34^. However, none of the previous studies has addressed the differences in early-stage innate immune cell responses between non-hospitalized (i.e., asymptomatic and mild patients) and hospitalized COVID-19 patients. Here, in contrast to the coordinated response in mild patients, our data revealed major innate immune cell dysregulations and impairment exclusively in hospitalized patients.

With few exceptions^7^, much less was known so far about protective anti-SARS-CoV-2 immune responses in non-hospitalized mild COVID-19 patients. Our current work now provides a first resource that systematically describes the evolving trajectory of highly coordinated early-stage protective immune responses in mild COVID-19 patients. Based on a sufficiently-powered sample size (63 mild participants) and on the prospective longitudinal nature of our study, we discovered the frequency of CD38^+^ among CD8 T cells and of mature DCs at an early disease stage (maximum three days post-PCR diagnosis) to be highly robust and even better predictors of ensuing humoral responses at three weeks than early plasmablast responses, pointing to a critical role of DC activation in coordinating very early antigen-specific T cell and later antibody responses. Thus, the immune signatures identified in our study bear the potential to be extrapolated to predict protective immune responses in vaccinated people early on. In fact, our discoveries are in line with a recent report showing that mRNA vaccination induces rapid abundant antigen-specific CD4 T-cell responses in SARS-CoV-2 naïve participants following the first dose^50^, which phenocopies our findings in mild COVID-19 patients, thus indicating that the discoveries of our study will have a more general impact for understanding and further dissecting SARS-CoV-2-specific immunity. In our cohort, we were unable to observe a clear protective immune signature in the peripheral blood of PCR-positive asymptomatic individuals throughout all cellular and humoral immune analyses. Although the sample size was relatively small in our asymptomatic category, the most likely explanation for this result is a more prominent role of a tissue-resident rather than a systemic immune response, where anti-viral immunity mainly occurs locally through pre-activated innate immune stimulation in epithelial cells of the upper airways, as recently shown in a pediatric cohort^51^.

## Data Availability

N.A.

## Acknowledgements

We first would like to acknowledge the active involvement of all the anonymous participants in the Predi-COVID cohort. We are also thankful for the excellent support of the recruitment team of the Predi-COVID cohort from CIEC of LIH. The Predi-COVID study is supported by the Luxembourg National Research Fund (FNR) (Predi-COVID, 14716273) and the André Losch Fondation. We also highly appreciate the expert support of the IBBL processing and biorepository teams. F.Q.H. was partially supported by FNR CORE programme grant (CORE/14/BM/8231540/GeDES), FNR AFR-RIKEN bilateral programme (TregBAR, 11228353, F.Q.H. and M.O.) and PRIDE programme grants (PRIDE/11012546/NEXTIMMUNE and PRIDE/10907093/CRITICS). C.H. was partially supported by the FNR fast-track call COVID-19/2020-1/14703957/COV-Immun.

## Author contributions

C.C. designed and performed the experiments, performed data analysis and drafted the manuscript. S.C., O.D., F.H., M.K. and W.A. performed parts of experiments. I.E. and A.F. coordinated the cohort recruitment. C.S., C.H., G.S., T.A. and K.G. collected samples and performed parts of experiments. A.C. and M.V. collected clinical data, quality control and data management. C.L.C. and I.M.K. analyzed the TCR repertoire data. A.S., A.D.S., A.C., F.B. and G.F. provided substantial insights and supervision into the project. F.Q.H. and M.O. conceived and oversaw the whole project and wrote and finalized the manuscript.

## Supplementary Figures

**Supplementary Figure 1.**
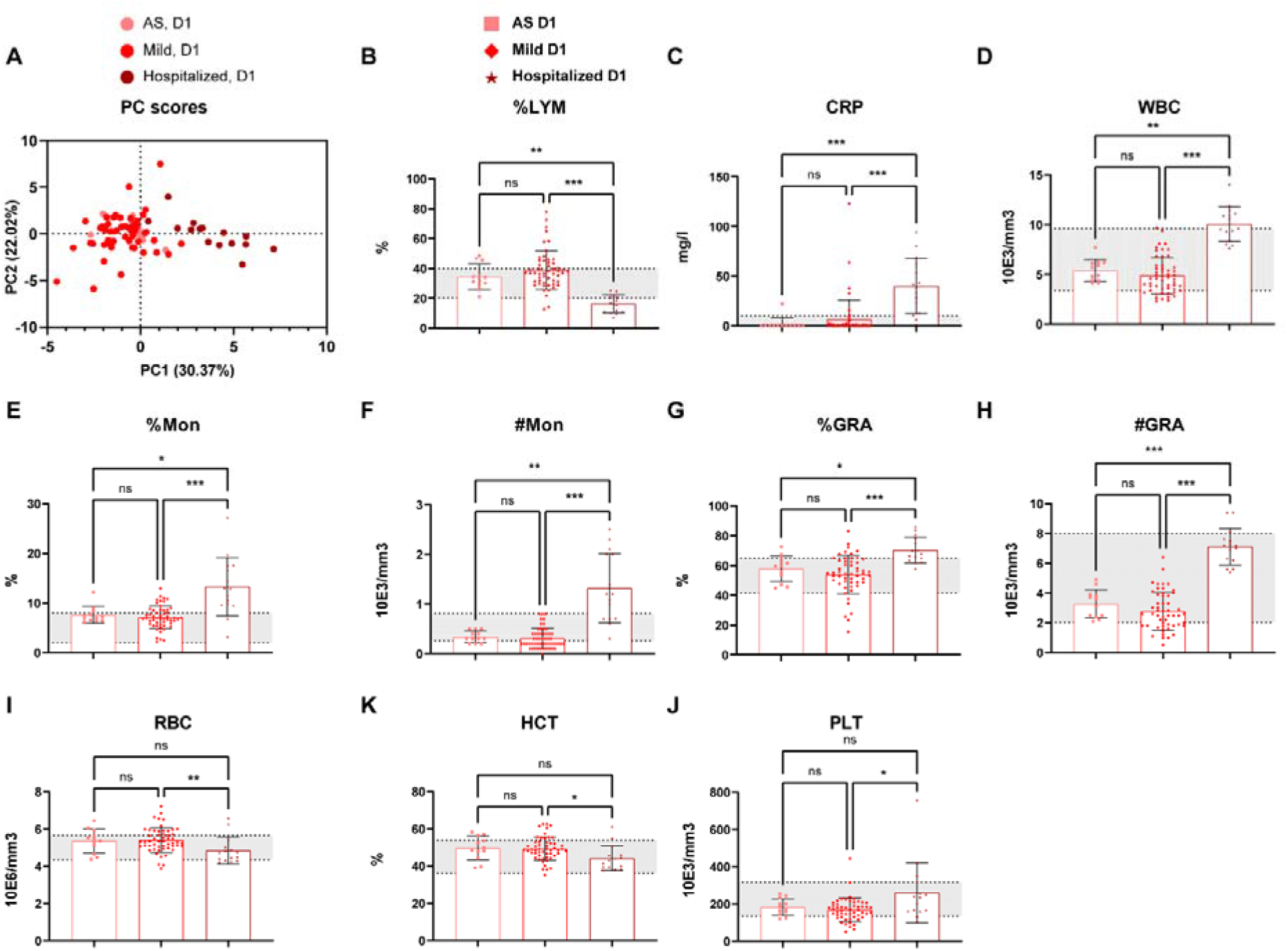
Whole-blood-count analysis of asymptomatic, mild and hospitalized patients at day 1 of inclusion. **A**, PCA plots of the samples from different patient groups at day 1 of inclusion based on 17 whole-blood-count parameters. AS, asymptomatic; HC, household controls; D1, day 1. **B, E, G**, the percentages of Lymphocytes (LYM, **B**), monocytes (Mon, **E**) and granulocytes (GRA, **G**). **C**, CRP (C-reactive protein) levels from different patient groups at day 1 of inclusion. **D, F, H, I, J**, Number of white blood cells (WBC, **D**), monocytes (Mon, **F**), granulocytes (GRA, **H**), red blood cells (RBC, **I**) and platelets (PLT, **J**) per ul (mm^3^). **K**, the hematocrit levels (%) from different patients groups at day 1 of inclusion. Data represent individual values; Mean± standard deviation (S.D.); P-value was determined by the Kruskal-Wallis (nonparametric) test and corrected using the Dunn’s multiple comparisons test. ns or unlabeled, not significant, *p<=0.05, **p<=0.01 and ***p<=0.001. Gray shading indicates the reported normal range for those different laboratory parameters.

**Supplementary Figure 2.**
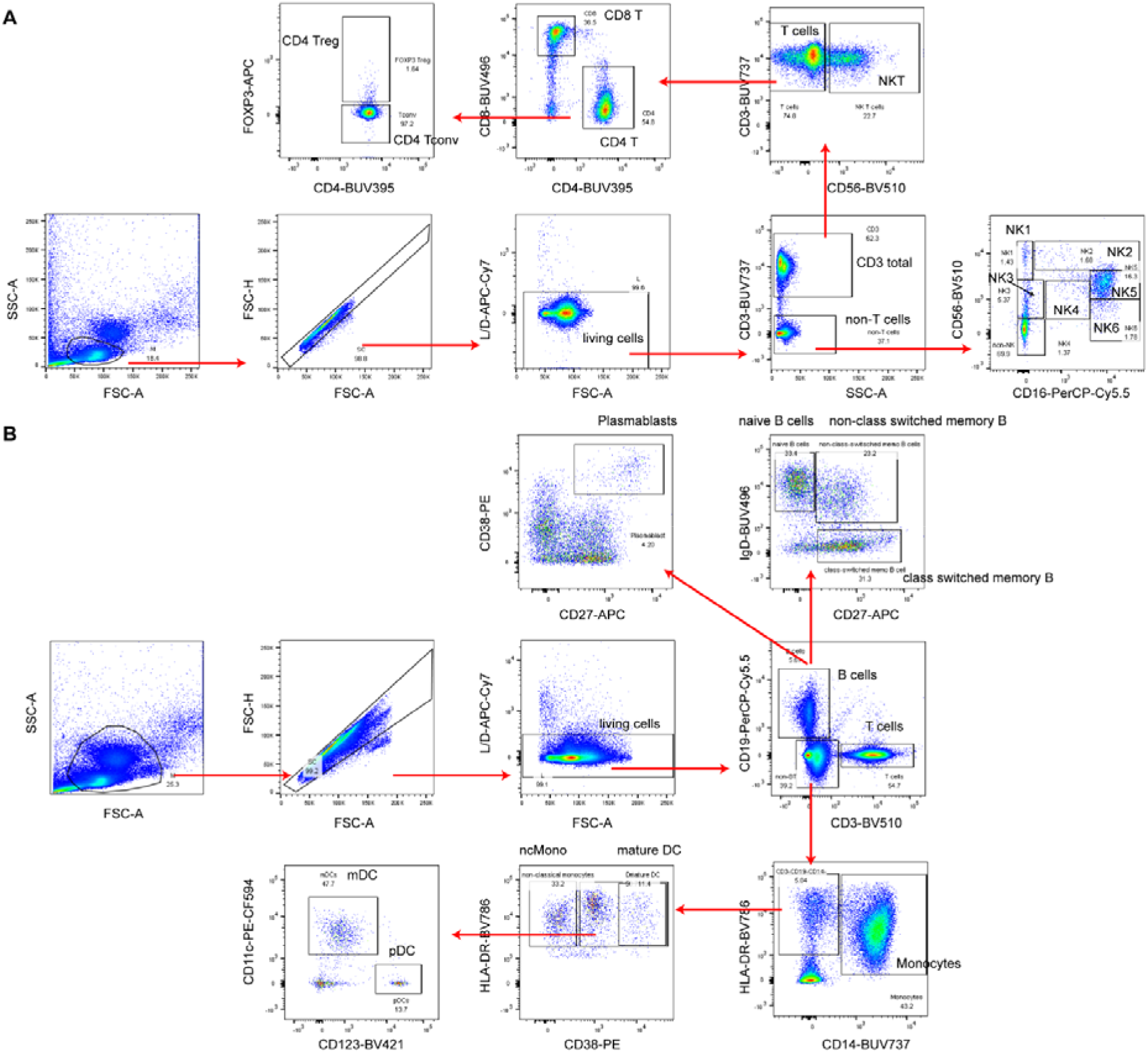
General gating strategy to identify different immune subsets analysed in our study. **A**, Lymphocyte gating strategy (not including B cells). Treg, FOXP3^+^CD4 T cells; Tconv, FOXP3^-^ CD4 T cells; NK, natural killer cells. **B,** Gating strategy to identify B cells, (non-)class-switched memory B cells, naïve B cells T cells, monocytes, DC, mDC, pDC, mature DC and plasmablasts. ncMono, non-classical monocytes; DC, Dendritic cells. Due to limited space, the functional markers or their combinations among each subset were not displayed here. It is also worthy to note that we used three different panels to perform a deep immunophenotyping analysis by flow cytometry. For some markers, different fluorochromes were used in different panels.

**Supplementary Figure 3.**
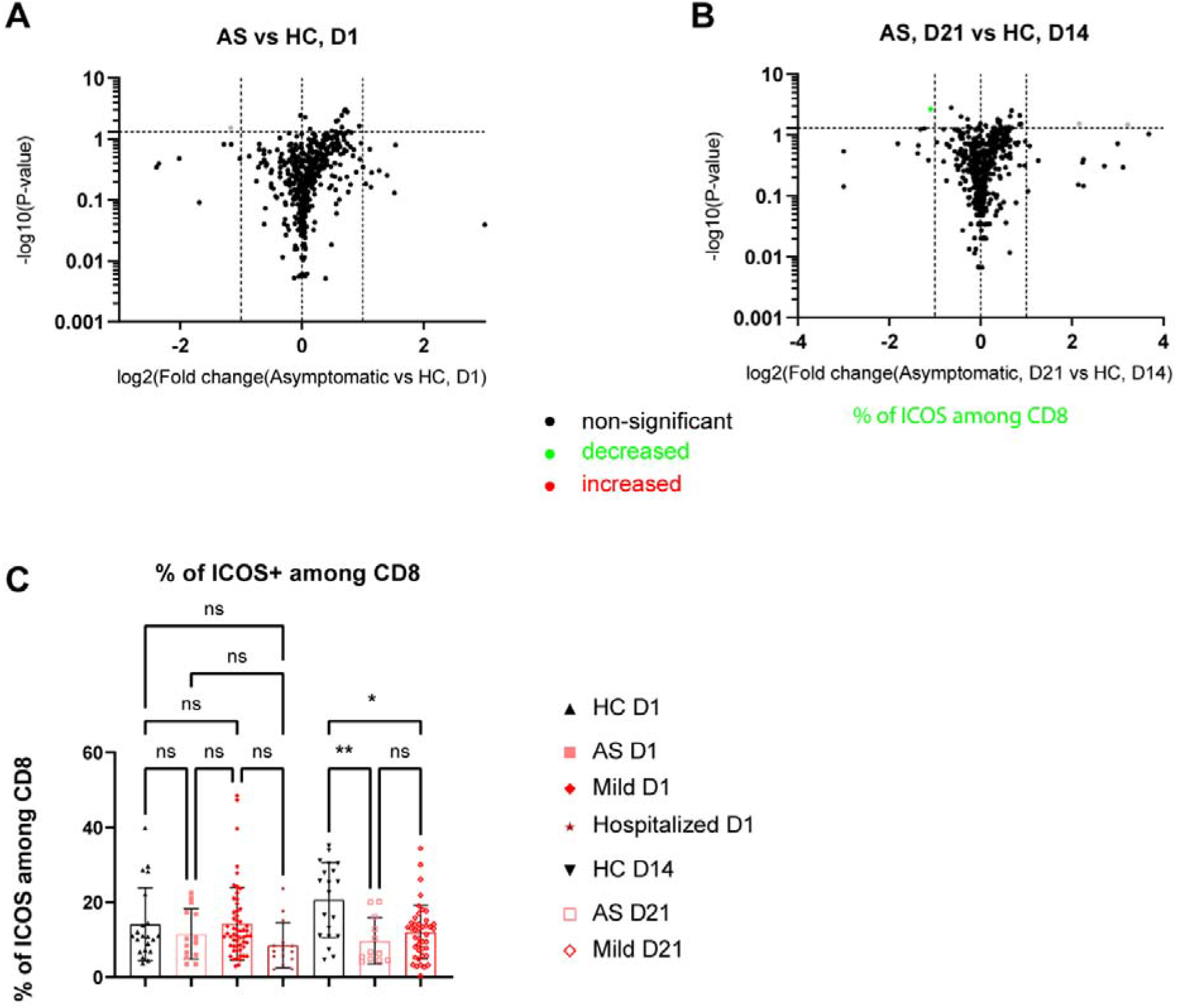
Comprehensive comparison of immune responses between asymptomatic patients and household controls. **A, B**, Volcano plots showing the immune responses in asymptomatic patients at day 1 (**A**) or day 21 (**B**) following inclusion. Significant increased or decreased subsets (p<=0.05 and change fold >=2) were marked in red or green, respectively. The gray dot represents the subset not showing a significant change after applying the Dunn’s multiple comparison test. **C**, Frequency of ICOS^+^ cells among CD8 T cells of different participant groups. AS, asymptomatic; HC, household controls; D1/D14/D21, day 1/day 14/day 21. Data represent individual values; Mean± standard deviation (S.D.); P-value was determined by the Kruskal-Wallis (nonparametric) test and corrected using the Dunn’s multiple comparisons test. ns or unlabeled, not significant, *p<=0.05, **p<=0.01 and ***p<=0.001.

**Supplementary Figure 4.**
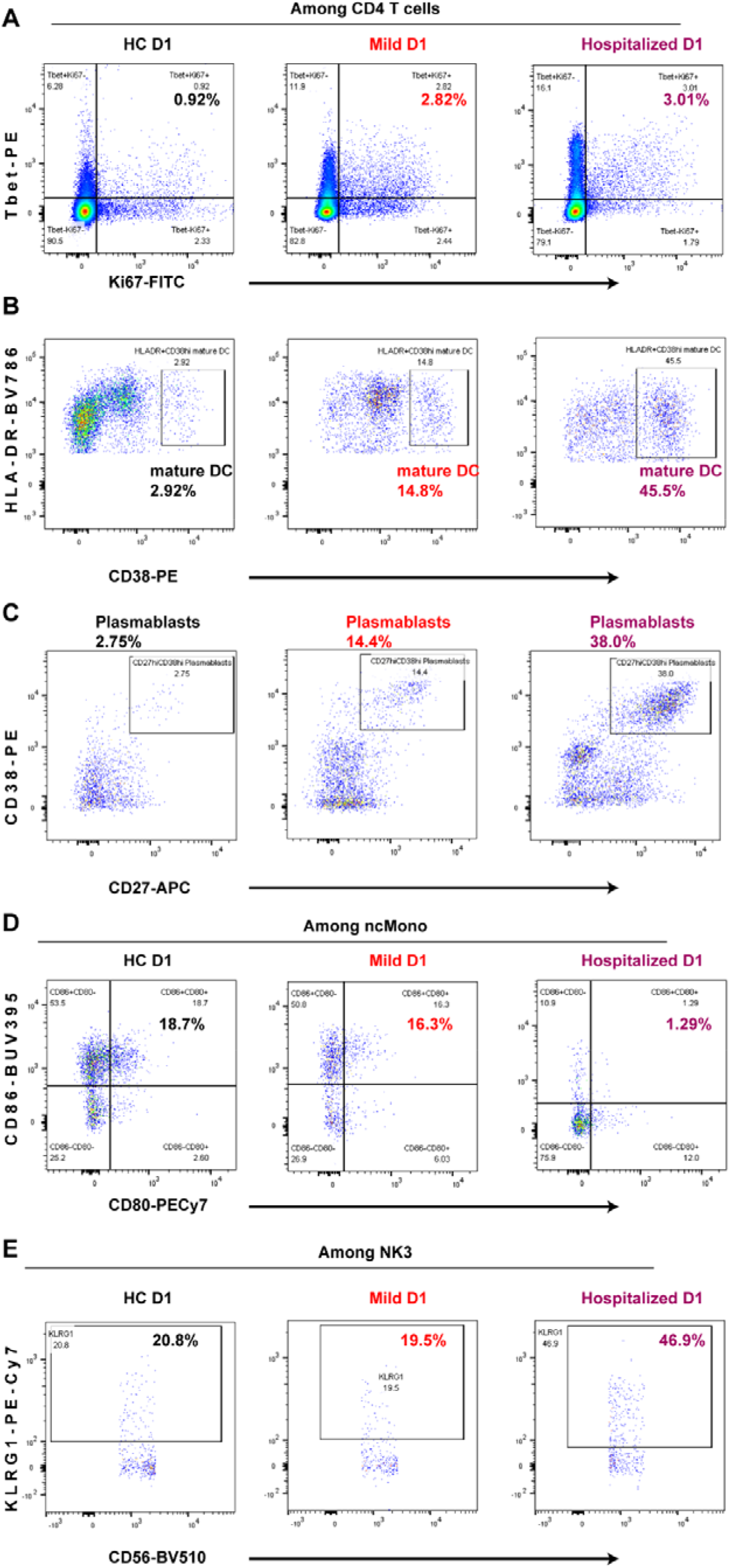
Extended representative flow cytometry plots in comparison of different groups at day 1 of inclusion. **A**, Representative flow-cytometry plots of the expression of Ki67 and Tbet among CD4 T cells from different participant groups. HC, household controls; D1, day 1. **B, C**, Representative flow-cytometry plots of the expression of CD38 and HLA-DR showing mature DC (**B**) or the expression of CD38 and CD27 showing plasmablasts (**C**) from different participant groups. **D**, Representative flow-cytometry plots of the expression CD86 and CD80 among non-classical monocytes (ncMono) from different groups. **E**, Representative flow-cytometry plots of the expression KLRG1 and CD56 among NK3 from different groups.

**Supplementary Figure 5.**
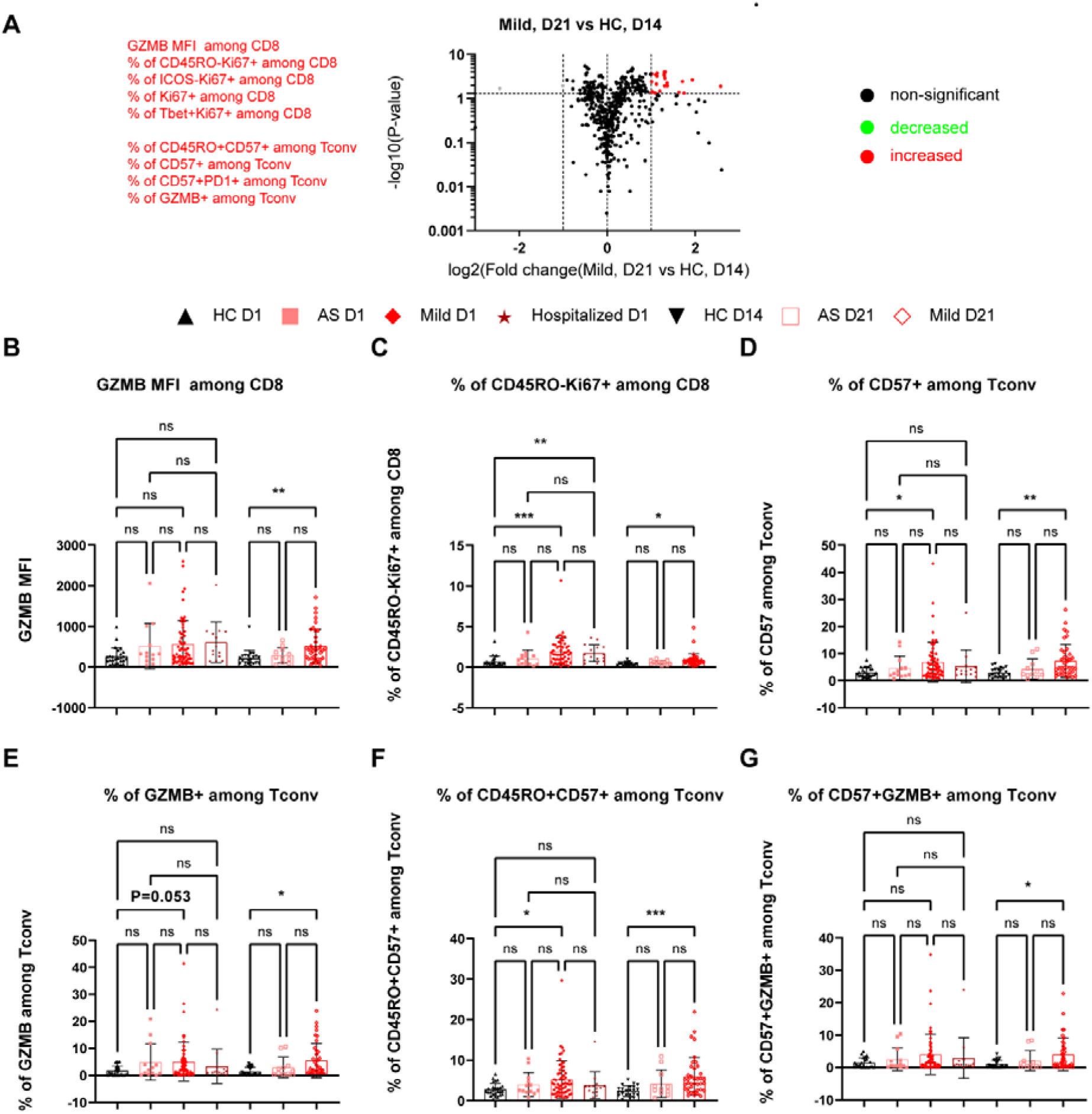
Comprehensive peripheral immune profiling of mild patients at day 21 following inclusion. **A**, Volcano plot showing responses of different immune subsets in mild patients at day 21 following inclusion relative to household controls (HC) at day 14 following inclusion. Significant increased or decreased subsets (p<=0.05 and change fold >=2) were marked in red or green, respectively. The gray dot represents the subset not showing a significant change after applying the Dunn’s multiple comparison test. **B**, GZMB MFI among CD8 T cells from different groups. AS, asymptomatic; HC, household controls; D1/D14/D21, day 1/day 14/day 21. **C**, Frequency of CD45RO^-^Ki67^+^ among CD8 T cells from different groups. **D, E, F, G**, Frequency of CD57^+^ (**D**), GZMB^+^ (**E**), CD45RO^+^CD57^+^ (**F**) or CD57^+^GZMB^+^ (**G**) among CD4 Tconv cells from different groups. Data represent individual values; Mean± standard deviation (S.D.); P-value was determined by the Kruskal-Wallis (nonparametric) test and corrected using the Dunn’s multiple comparisons test. ns or unlabeled, not significant, *p<=0.05, **p<=0.01 and ***p<=0.001.

**Supplementary Figure 6.**
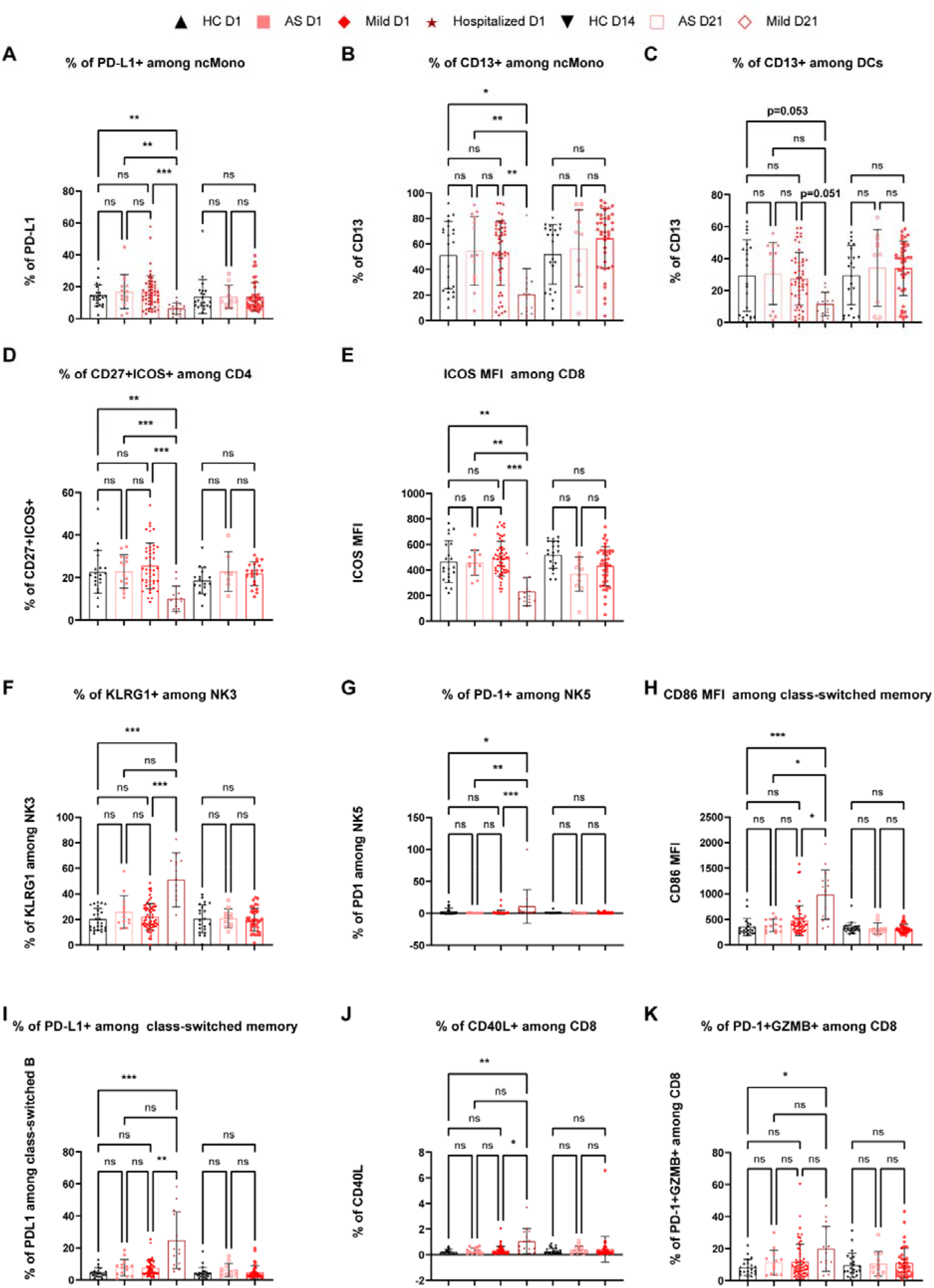
Extended analysis of early-stage immune features characterizing mild patents from hospitalized ones. **A, B**, Frequency of cells expressing PD-L1 (**A**) or CD13 (**B**) among ncMono from different participant groups. AS, asymptomatic; HC, household controls; D1/D14/D21, day 1/day 14/day 21. **C**, Frequency of CD13^+^ among HLA-DR^+^CD38^+^ DCs. **D, E,** Frequency of CD27^+^ICOS^+^ among CD4 cells (**D**) or ICOS MFI (**E**) among CD8 T cells. **F, G**, Frequency of KLRG1^+^ cells among NK3 (**F**) or frequency of PD-1^+^ cells among NK5 (**G**). **H, I**, CD86 MFI (**H**) or the frequency of PD-L1^+^ cells (**I**) among class-switched memory B cells. **J, K**, Frequency of CD40L^+^ cells (**J**) or frequency of PD-1^+^GZMB^+^ cells (**K**) among CD8 T cells. Data represent individual values; Mean± standard deviation (S.D.); P-value was determined by the Kruskal-Wallis (nonparametric) test and corrected using the Dunn’s multiple comparisons test. ns or unlabeled, not significant, *p<=0.05, **p<=0.01 and ***p<=0.001.

**Supplementary Figure 7.**
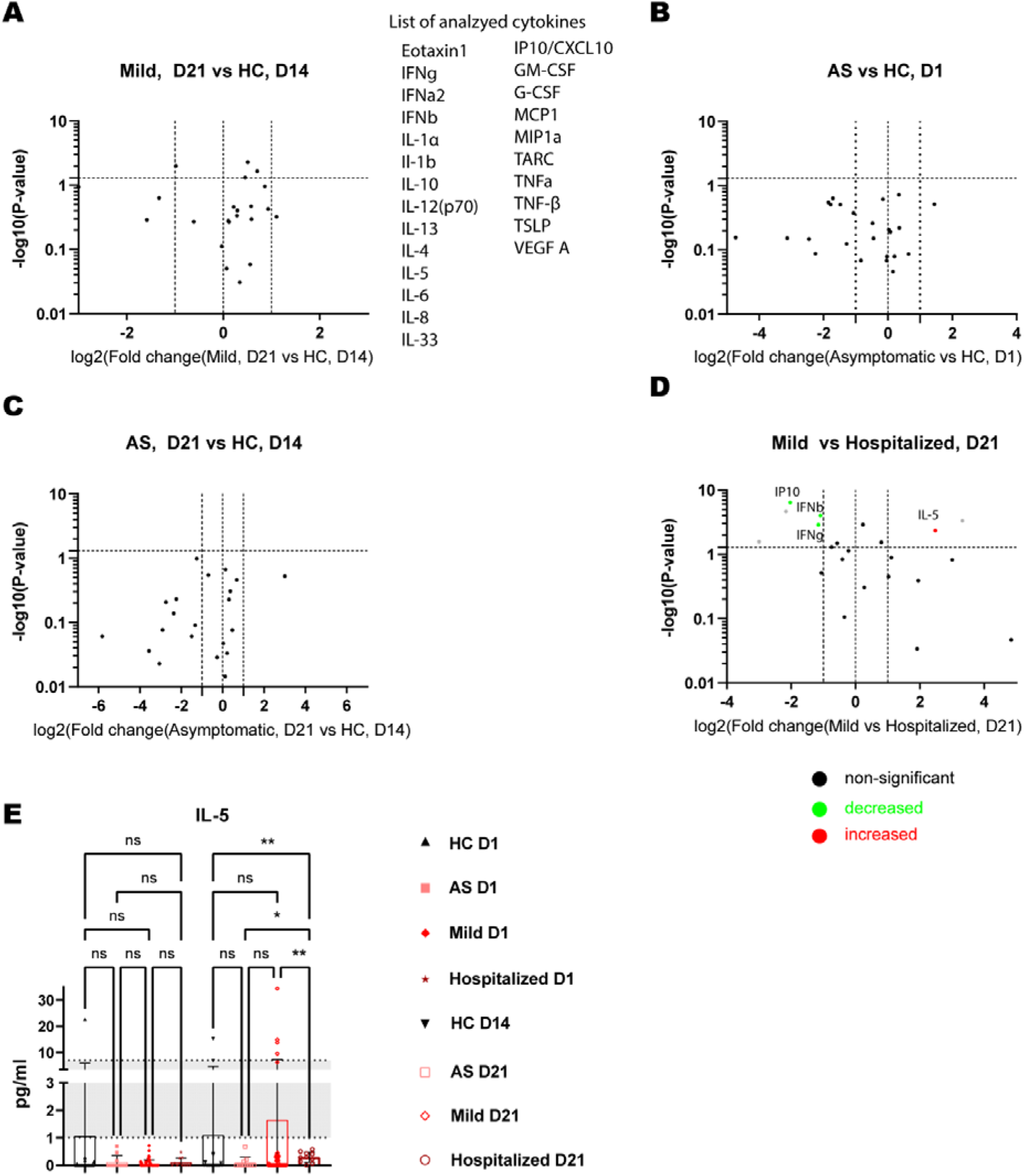
Extended comparison of serological cytokine/chemokine responses between different patient groups at day 1 or day 21 following inclusion. **A**, Volcano plot showing cytokine/chemokine responses of mild patients at day 21 versus household controls (HC) at day 14 following inclusion. **B**, Volcano plot showing cytokine/chemokine responses of asymptomatic patients versus household controls (HC) at day 1 of inclusion. **C**, Volcano plot showing cytokine/chemokine responses of asymptomatic (AS) patients at day 21 versus household controls (HC) at day 14 following inclusion. **D**, Volcano plot showing cytokine/chemokine responses of mild patients versus hospitalized patients at day 21 of inclusion. The gray dot represents the analytes showing a significant change but displaying values lower than the reported normal physiological levels even in control groups. **E**, Scatter dot plots of serological levels of IL-5 of different participant groups in the cohort. Gray shading indicates the reported normal range for IL-5. AS, asymptomatic; HC, household controls; D1/D14/D21, day 1/day 14/day 21. Data represent individual values; Mean± standard deviation (S.D.); P-value was determined by the Kruskal-Wallis (nonparametric) test and corrected using the Dunn’s multiple comparisons test. ns or unlabeled, not significant, *p<=0.05, **p<=0.01 and ***p<=0.001.

## Materials and Methods

### Cohort design

Predi-COVID is a prospective longitudinal cohort study composed of individuals older than 18 years of age with a positive PCR test for SARS-CoV-2 in Luxembourg. Blood samples were collected by a nurse at the latest 3 days post clinical PCR diagnosis (baseline, as day 1) at home for asymptomatic and mild participants. For hospitalized patients, except for two of them (sampled 5 or 6 days post hospital arrival), the remaining 13 patients were all sampled at the latest 3 days after hospitalization. A follow-up visit was organized 3 weeks (day 21) later.

The Predi-COVID-H sub-study is a prospective longitudinal cohort study composed of household members of a Predi-COVID participant as controls. Biological samples were collected at the same time as for the Predi-COVID participant sharing the house (baseline, as day 1) and 2 weeks later, at day 14. More details on the study design have been described ^16^.

### Blood sampling and PBMC isolation

Samples were collected from confirmed SARS-COV-2 positive patients and household controls by trained nurses from the LIH-CIEC. Blood samples were collected in CAT, K_2_EDTA and CPT (all from BD, Erembodegem, Belgium) by the standard phlebotomy procedure. Blood samples were transported daily to centralized processing laboratory (IBBL) at ambient temperature.

CAT tubes were centrifuged for 10 min at 2000 x g room temperature (RT). Serum upper layer was sterile aliquoted and stored at −80 °C. Prior to centrifugation, 200 µl from the K_2_EDTA was transferred into 0.5 ml microcentrifuge tube for complete blood count (CBC) on ABX Micros CRP200 (Horiba, Japan). The K_2_EDTA tube was centrifuged for 20 min at 2000 x g, 4 °C. Plasma upper layer and buffy-coat were aliquoted and stored at −80 °C. The CPT tubes were centrifuged for 20 min at 1800 x g, RT. The collected PBMCs were washed twice in PBS and counted using a Cellometer (Nexcelom, UK). Fresh PBMCs were partly used for direct flow cytometry and partly cryopreserved in CryoStor CS10 (Biolife solutions, USA) by controlled-rate freezing using Mr. Forsty (Nalgene, USA), followed by a long-term storage in liquid nitrogen.

### Flow cytometry

For each panel staining, 1×10^6^ isolated fresh PBMCs, rather than frozen PBMCs, were used since cryopreservation affects several relevant markers^52^. The cells were resuspended in 50 ul of Brilliant stain buffer (BD, 563794) containing 2.5 ul of Fc blocking antibodies (BD, 564765) and incubated for 15 min. The suspension was then mixed with 50 ul of the respective 2x concentrated mastermix for the surface staining. The staining concentration and fluorochromes of the different markers are specified in **Supplementary Table 2**. After 30 min of incubation in the dark at 4 °C, the cells were washed three times with FCM buffer (flow cytometry [FCM] staining buffer, Ca^2+^- and Mg^2+^-free PBS + 2% heat-inactivated FBS) (5 min, 300 x g). Following the final washing step, the stained PBMCs were fixed in 200 ul of 4% PFA (ThermoFisher Scientific, 28906) and incubated at RT for 30 min in the dark. After the PFA fixation, the PBMCs were washed once in FCM buffer (5 min, 400 x g) and resuspended in 200 ul fixation buffer from the True-Nuclear Transcription Factor Buffer Set (Biolegend, 424401). After 1 h of incubation in the dark at RT, the cells were centrifuged down (5 min, 400 x g) and resuspened in 200 ul FCM buffer and left at 4 °C overnight.

In the next morning the cells were resuspended in permeabilisation buffer (Biolegend, 424401) containing 2.5 ul of Fc blocking antibodies (BD, 564765) and incubated for 15 min at RT. After the intracellular blocking step, the cells were resuspended in 100 ul permeabilisation buffer containing the antibodies for the intracellular staining (**Supplementary Table 2**). After 30 min of incubation at RT in the dark, the cells were washed 3 times with permeabilisation buffer (5 min, 400 x g) and resuspended in 100 ul FCM buffer to proceed to the acquisition on a BD LSRFortessa^TM^ analyzer. To ensure a consistent acquisition of all the markers over the whole duration of the study, the application settings of the instrument were saved during the first acquisition and applied to all the following samples of the cohort. The data was analyzed using FlowJo v10.5.6.

### Determination of cytokine and chemokine levels by MSD assay

24 cytokines, chemokines or growth factors (eotaxin-1, G-CSF, GM-CSF, IFN-α2a, IFN-β, IFN-γ, IL-1α, IL-1β, IL-4, IL-5, IL-6, IL-8, IL-10, IL12p70, IL-13, IL-33, IP10, MCP-1, MIP-1α, TARC, TNF-α, TNF-β, TSLP, VEGFA) were measured in patient sera using a multiplex assay (U-Plex Biomarker Group 1 (hu) assays from MSD Kit catalog Number K15067L-1). The samples were undiluted. The assay was performed according to the manufacturer’s instructions. Data were recorded and analyzed on a MESO QuickPlex SQ 120 instrument.

### Serological detection of IgG against SARS-CoV-2 by MSD assay

V-Plex COVID-19 Coronavirus Panel1 serology kits from MSD (reference K15362U) were used to detect the presence of IgG antibodies to SARS-CoV-2-Spike (S), SARS-CoV-2 Nucleocapsid (N), SARS-CoV-2-S NTD (NTD) and SARS-CoV-2-S RBD (RBD) in diluted sera (1/500) according to the manufacturer’s instructions. The plate was read on an MSD instrument, which measures the light emitted from the MSD SULFO-TAG. To determine the cutoff values for positivity for SARS-CoV-2, we measured 35 patients in another cohort from Central Hospital of Luxembourg (PCR positive and >15 days symptom onset) and negative sera before the pandemic from 2019 stored in Luxembourg National Laboratory (LNS). The values of IgG for severe ones from that cohort as defined positive controls have already been displayed in Fig. 1. Of note, the hospitalized samples (both at day 1 and day 21 of inclusion) in this cohort were measured using lots of plates different from the other groups and the corresponding positive thresholds of those plates were calculated accordingly. To guarantee the comparability of positive percentages between the two batches, the choice of the cutoffs aimed for a similar sensitivity and specificity between the two batches of plates (the AUC analysis was done by GraphPad Prism 9.0).

### Determination of neutralization antibody capacity by MSD assay

Multiplex assays for the detection of neutralizing antibodies against SARS-CoV-2 (SARS-CoV-2-Spike and SARS-CoV-2 S RBD) were done on patient sera using the MSD COVID-19 ACE2 Neutralization Kits from MSD (Panel 1 reference K15375U) according to the manufacturer’s instructions. The samples were diluted 50 times for the neutralization assay. Data were recorded on a MESO QuickPlex SQ 120 instrument, which measures the light emitted from the MSD SULFO-TAG. Results were reported as percent inhibition calculated using the equation below. % Inhibition was calculated using the following equation: (1-Average Sample ECL Signal/Average ECL signal of calibrator) *100.

### TCR repertoire analysis

Immunosequencing of the CDR3 regions of human TCRβ chains was performed using the ImmunoSEQ→ Assay (Adaptive Biotechnologies, Seattle, WA). Extracted genomic DNA from ∼5e6 cryopreserved PBMCs was amplified in a bias-controlled multiplex PCR, followed by high-throughput sequencing. Sequences were collapsed and filtered in order to identify and quantitate the absolute abundance of each unique TCRβ CDR3 region for further analysis as previously described^53^.

Clonality was defined as 1-normalized Shannon’s Entropy and was calculated on productive rearrangements. Clonality values approaching 0 indicate a very even distribution of frequencies, whereas values approaching 1 indicate an increasingly asymmetric distribution in which a few clones are present at high frequencies. Clonal breadth and depth of SARS-CoV2-associated TCRβ sequences were calculated as previously described^28^, using a set of sequences described elsewhere^29^. Briefly, breadth is calculated as the proportion of unique annotated SARS-CoV-2 specific rearrangements out of the total number of unique productive rearrangements, while depth accounts for the extent of the expansion of those clonal lineages in the repertoire.

### Ethics statements

All collections were performed with approval from relevant ethic organizations. Informed consent was obtained from each participant prior to collection. The blood sampling was performed by nurses from Clinical and Epidemiological Investigation Centre (CIEC) of LIH.

### Statistical analysis

Both PCA and volcano plots were visualized using GraphPad Prism 9.0. Correlation analysis was based on either Spearman or Pearson correlation as indicated in the corresponding figures. We only displayed the top-ranked highly correlated results if the correlation coefficient was ranked in the top positions among 484 correlations, calculated between antibody levels, cytokines or TCR breadth and any of the 484 subsets. The corresponding P-values from correlation analysis were based on a two-tailed analysis using GraphPad Prism 9.0. P-value from each scatter dot plots was determined by the Kruskal-Wallis (nonparametric) test and corrected using the Dunn’s multiple comparisons test from GraphPad Prism 9.0. In addition to each individual value, data from each group were presented as mean± standard deviation (S.D.). ns or unlabeled, not significant, *p<=0.05, **p<=0.01 and ***p<=0.001.

## Supplementary Tables

**Supplementary Table 1.** Demographics of our longitudinal COVID-19 cohort.

| Clinical characteristics of the cohort | All patient s, D1 | Asymp to matic, D1 | Mild, D1 | Hospit alized, D1 | Househo ld controls, D1 | All patient s, D21 | Asymp to matic , D21 | Mild, D21 | Househo ld Controls , D14 |
| --- | --- | --- | --- | --- | --- | --- | --- | --- | --- |
| median (IQR) |  |  |  |  |  |  |  |  |  |
| Age (years) | 44 (30-53) | 44 (24.5-55.5) | 38 (28.5-48) | 57 (49-61) | 33.5 (26-40.75) | 40.5 (30.8-48.5) | 44 (29-53) | 40 (31-48) | 33.5 (27-42.25) |
| BMI | 25.7 (23.5-28.6) | 24.8 (22.9-26.6) | 25.8 (23.5-29.4) | 26.64 (24.9-29.1) |  | 25.9 (23.9-27.9) | 24.8 (22.3-26.1) | 26.3 (24.4-28.7) |  |
| concerned n (percentage among the given category) |  |  |  |  |  |  |  |  |  |
| Male (%) | 54 (63.5%) | 9 (69.2%) | 39 (63.9%) | 8 (57.1%) | 7 (30.4%) | 33 (66%) | 8 (72.7%) | 25 (64.1%) | 7 (35%) |
| Smoking (%) | 14 (18.7%) | 3 (23.1%) | 11 (18.1%) |  |  | 9 (18%) | 2 (18.2%) | 7 (18%) |  |
| Former smoker (%) | 16 (21.4%) | 2 (15.4%) | 14 (23%) |  |  | 12 (24%) | 2 (18.2%) | 10 (25.7%) |  |
| Whole Blood Count parameters: median (IQR) |  |  |  |  |  |  |  |  |  |
| WBC, white blood cell count (10E3/uL) | 5.25 (3.9-7.625) | 5.3 (4.4-6.1) | 4.6 (3.5-5.7) | 9.6 (8.85-11.05) |  |  |  |  |  |
| #Lymphocyte, LYM (10E3/uL) | 1.65 (1.2-2.1) | 1.8 (1.4-2.1) | 1.6 (1.2-2.1) | 1.6 (1-2.1) |  |  |  |  |  |
| %Lymphocyte, LYM (%) | 34.05 (25.575-40.75) | 33 (29.8-37.7) | 36.9 (31-43.9) | 16.8 (10.95-21.3) |  |  |  |  |  |
| #Monocyte, MON (10E3/uL) | 0.3 (0.2-0.6) | 0.3 (0.28-0.43) | 0.2 (0.2-0.4) | 1.2 (0.7-2) |  |  |  |  |  |
| %Monocyte, MON (%) | 7.3 (6.4-9.65) | 7.15 (6.5-7.9) | 6.9 (5.8-8.5) | 12.8 (9.7-17.1) |  |  |  |  |  |
| #Granulocyte, GRA (10E3/uL) | 3.05 (2.175-4.6) | 3.35 (2.5-3.8) | 2.6 (1.8-3.8) | 7.2 (6.25-7.55) |  |  |  |  |  |
| %Granulocyte, GRA (%) | 58.1 (50.9-65.2) | 59.7 (52.1-63) | 54.7 (48.7-62.7) | 67.8 (64.15-77.05) |  |  |  |  |  |
| Red Blood Cell, RBC (10E6/uL) | 5.265 (4.865-5.7125) | 5.41 (5.0-5.7) | 5.31 (5.1-5.9) | 4.73 (4.335-4.945) |  |  |  |  |  |
| Hemoglobin, HGB (g/dl) | 15.5 (14.1-17.025) | 15.6 (15.1-17.4) | 15.8 (14.7-17) | 13.9 (13.05-14.85) |  |  |  |  |  |
| **Hematocrit, HCT (%)** | 48.25<br>(44.05-52.65) | 49.1<br>(46.9-54.4) | 48.8<br>(45.6-52.8) | 42.9<br>(39.5-45.7) |  |  |  |  |  |
| **Platelet, PLT (10E3/uL)** | 175<br>(144-212) | 179.5<br>(154.8-209.3) | 167<br>(130-199) | 235<br>(163-286.5) |  |  |  |  |  |
| **Mean Cell Volume, MCV (um3)** | 92 (89-94) | 92.5<br>(89.8-95) | 92 (89-94) | 91 (90-93) |  |  |  |  |  |
| **Mean Cell Hemoglobin, MCH (pg)** | 29.35<br>(28.5-30.85) | 29.35<br>(28.5-30.3) | 28.9<br>(28.5-30.7) | 30.4<br>(29.9-31) |  |  |  |  |  |
| **Mean Cell Hemoglobin Concentration, MCHC (g/dL)** | 32.1<br>(31.6-32.6) | 31.9<br>(31.8-32.4) | 32<br>(31.4-32.4) | 33.3<br>(33-33.5) |  |  |  |  |  |
| **Red Cell Distribution Width, RDW (%)** | 14.35<br>(14-14.7) | 14.45<br>(14.1-14.6) | 14.4<br>(14.1-14.8) | 14.1<br>(13.95-14.55) |  |  |  |  |  |
| **Mean Platelet Volume, MPV (um3)** | 8.4 (8-8.8) | 8.35<br>(8.1-9.0) | 8.4 (8-8.7) | 8.3 (8-8.6) |  |  |  |  |  |
| **C-reactive protein, CRP (mg/L)** | 0.65 (0-10.075) | 0 (0-0.03) | 0.4 (0-1.6) | 38<br>(18.4-58) |  |  |  |  |  |
| **Comorbidity: concerned n (the percentage among the given category)<sup>s</sup>** |  |  |  |  |  |  |  |  |  |
| **Hypertension** | 8(10.7%) | 2(15.4%) | 6(9.9%) |  |  | 6(12%) | 1(9.1%) | 5<br>(12.9%) | NA* |
| **Chronic heart disease, including congenital heart disease (except hypertension)** | 5(6.7%) | 1(7.7%) | 4(6.6%) |  |  | 2 (4%) | 1(9.1%) | 1(2.6%) | NA |
| **Chronic lung disease (except asthma)** | 0 (0%) | 0 (0%) | 0 (0%) |  |  | 0 (0%) | 0 (0%) | 0 (0%) | NA |
| **Asthma (clinical diagnosis made)** | 3 (4%) | 0 (0%) | 3 (5%) |  |  | 0 (0%) | 0 (0%) | 0 (0%) | NA |
| **Chronic kidney disease** | 0 (0%) | 0 (0%) | 0 (0%) |  |  | 0 (0%) | 0 (0%) | 0 (0%) | NA |
| **Renal failure requiring dialysis** | 0 (0%) | 0 (0%) | 0 (0%) |  |  | 0 (0%) | 0 (0%) | 0 (0%) | NA |
| **Moderate or severe liver disease** | 1(1.4%) | 0 (0%) | 1 (1.7%) |  |  | 1 (2%) | 0 (0%) | 1(2.6%) | NA |
| **Mild liver disease** | 1(1.4%) | 1 (7.7%) | 0 (0%) |  |  | 1 (2%) | 1 (9.1%) | 0 (0%) | NA |
| **Chronic neurological disorder** | 1(1.4%) | 0 (0%) | 1 (1.7%) |  |  | 0 (0%) | 0 (0%) | 0 (0%) | NA |
| **Cancer** | 0 (0%) | 0 (0%) | 0 (0%) |  |  | 0 (0%) | 0 (0%) | 0 (0%) | NA |
| **Chronic hematologic disease** | 3 (4%) | 0 (0%) | 3 (5%) |  |  | 1 (2%) | 0 (0%) | 1(2.6%) | NA |
| **HIV / AIDS** | 0 (0%) | 0 (0%) | 0 (0%) |  |  | 0 (0%) | 0 (0%) | 0 (0%) | NA |
| **Obesity (clinical diagnosis made)** | 4(5.4%) | 0 (0%) | 4 (6.6%) |  |  | 4 (8%) | 0 (0%) | 4(10.3%) | NA |
| **Diabetes with associated complications** | 0 (0%) | 0 (0%) | 0 (0%) |  |  | 0 (0%) | 0 (0%) | 0 (0%) | NA |
| **Uncomplicated diabetes** | 5<br>(6.7%) | 0 (0%) | 5 (8.2%) |  |  | 2 (4%) | 0 (0%) | 2(5.2%) | NA |
| **Rheumatologic disease** | 1<br>(1.4%) | 0 (0%) | 1 (1.7%) |  |  | 1 (2%) | 0 (0%) | 1(2.6%) | NA |
| **Malnutrition** | 1(1.4%) | 0 (0%) | 1 (1.7%) |  |  | 1 (2%) | 0 (0%) | 1(2.6%) | NA |
| **Chronic obstructive pulmonary disease (COPD)** | 1(1.4%) | 0 (0%) | 1 (1.7%) |  |  | 0 (0%) | 0 (0%) | 0 (0%) | NA |
| **Other notable diseases or risk factors** | 12(16%) | 1 (7.7%) | 11(18.1%) |  |  | 8 (16%) | 0 (0%) | 8(20.6%) | NA |

§, the total number of patients only include asymptomatic and mild patients, but not hospitalized patients.
\*, NA or empty, no information available.
IQR: 25% percentile-75% percentile.
n, the number of participants in the given category.

**Supplementary Table 2.** List of antibodies used in this study.

| Marker and Fluorochrome | Supplier | Reference | Dilution |
| --- | --- | --- | --- |
| Fc Block | BD | 564765 | 2.5ul/1M cells |
| CD4 BUV395 | BD | 563550 | 1 :200 |
| CD8 BUV496 | BD | 612942 | 1 :200 |
| CD3 BUV737 | BD | 741822 | 1 :200 |
| CD56 BV510 | BD | 740171 | 1 :50 |
| Tim3 BV786 | BD | 742857 | 1 :50 |
| CD57 FITC | BD | 561906 | 1 :100 |
| CD16 BB700 | BD | 746199 | 1 :50 |
| GZMB PE (intracellular)<sup>§</sup> | BD | 561142 | 1 :20 |
| CD45RO PE-CF594 | BD | 562299 | 1 :50 |
| CD38 BV510* | BD | 563251 | 1 :25 |
| CXCR5 BV711 | BD | 740737 | 1 :200 |
| HLA-DR BV786 | BD | 564041 | 1 :50 |
| Ki67 AF588 (intracellular) | BD | 561165 | 1 :50 |
| CD27 BB700 | BD | 566449 | 1 :50 |
| CD40L PE-Cy5 | BD | 555701 | 1 :50 |
| CD86 BUV395 | BD | 740305 | 1 :25 |
| IgD BUV 496 | BD | 741170 | 1 :200 |
| CD14 BUV737 | BD | 612763 | 1 :100 |
| CD123 BV421 | BD | 563362 | 1 :50 |
| CD3 BV510 | BD | 564713 | 1 :100 |
| CD40 BV605 | BD | 740410 | 1 :200 |
| CD13 BV711 | BD | 740772 | 1 :25 |
| PD-L1 BB515 | BD | 564554 | 1 :50 |
| CD19 BB700 | BD | 566411 | 1 :200 |
| CD38 PE | BD | 560981 | 1 :25 |
| CD11c PE-CF594 | BD | 562393 | 1 :50 |
| CD80 PE-CY7 | BD | 561135 | 1 :25 |
| CD27 APC | BD | 561786 | 1 :50 |
| ICOS (CD278) BV605 | BioLegend | 313538 | 1 :100 |
| KLRG1 PE-Cy7 | BioLegend | 368614 | 1 :50 |
| FOXP3 APC (intracellular) | BioLegend | 320114 | 1 :20 |
| CCR7 (CD197) BV421 | BioLegend | 353208 | 1 :50 |
| CD45RA BV421 | BioLegend | 304130 | 1 :50 |
| PD-1 (CD279) BV605 | BioLegend | 329924 | 1 :50 |
| CD127 BV711 | BioLegend | 351328 | 1 :100 |
| T-bet PE (intracellular) | BioLegend | 644810 | 1 :50 |
| CD194 APC | BioLegend | 359408 | 1 :100 |
| Eomes PE-Cy7 (intracellular) | ThermoFischer Scientific | 25-4877-42 | 1 :20 |
| True-Nuclear kit | BioLegend | 424401 |  |
| Ultracomp Comp Beads | ThermoFischer Scientific | 01-2222-42 |  |

§, intracellular, indicating an intracellular staining protocol was applied; otherwise, cell surface staining.
\*, different fluorochromes were used for some markers among different panels.

## References

1 Mathieu, E. et al. A global database of COVID-19 vaccinations. Nature Human Behaviour 5, 947–953, doi:10.1038/s41562-021-01122-8 (2021).

2 Kuri-Cervantes, L. et al. Comprehensive mapping of immune perturbations associated with severe COVID-19. Science Immunology 5, eabd7114, doi:10.1126/sciimmunol.abd7114 (2020).

3 Mathew, D. et al. Deep immune profiling of COVID-19 patients reveals distinct immunotypes with therapeutic implications. Science 369, eabc8511, doi:10.1126/science.abc8511 (2020).

4 Arunachalam, P. S. et al. Systems biological assessment of immunity to mild versus severe COVID-19 infection in humans. Science 369, 1210–1220, doi:10.1126/science.abc6261 (2020).

5 De Biasi, S. et al. Marked T cell activation, senescence, exhaustion and skewing towards TH17 in patients with COVID-19 pneumonia. Nat Commun 11, 3434, doi:10.1038/s41467-020-17292-4 (2020).

6 Zheng, M. et al. Functional exhaustion of antiviral lymphocytes in COVID-19 patients. Cellular & Molecular Immunology 17, 533–535, doi:10.1038/s41423-020-0402-2 (2020).

7 Bergamaschi, L. et al. Longitudinal analysis reveals that delayed bystander CD8+ T cell activation and early immune pathology distinguish severe COVID-19 from mild disease. Immunity, doi:10.1016/j.immuni.2021.05.010 (2021).

8 Wilk, A. J. et al. A single-cell atlas of the peripheral immune response in patients with severe COVID-19. Nat Med 26, 1070–1076, doi:10.1038/s41591-020-0944-y (2020).

9 Saichi, M. et al. Single-cell RNA sequencing of blood antigen-presenting cells in severe COVID-19 reveals multi-process defects in antiviral immunity. Nature Cell Biology 23, 538–551, doi:10.1038/s41556-021-00681-2 (2021).

10 Ren, X. et al. COVID-19 immune features revealed by a large-scale single-cell transcriptome atlas. Cell 184, 1895–1913 e1819, doi:10.1016/j.cell.2021.01.053 (2021).

11 Long, Q.-X. et al. Clinical and immunological assessment of asymptomatic SARS-CoV-2 infections. Nature Medicine 26, 1200–1204, doi:10.1038/s41591-020-0965-6 (2020).

12 Sekine, T. et al. Robust T Cell Immunity in Convalescent Individuals with Asymptomatic or Mild COVID-19. Cell 183, 158–168.e114, doi:https://doi.org/10.1016/j.cell.2020.08.017 (2020).

13 Le Bert, N. et al. SARS-CoV-2-specific T cell immunity in cases of COVID-19 and SARS, and uninfected controls. Nature 584, 457–462, doi:10.1038/s41586-020-2550-z (2020).

14 Delhalle, S., Bode, S. F. N., Balling, R., Ollert, M. & He, F. Q. A roadmap towards personalized immunology. npj Systems Biology and Applications 4, 9, doi:10.1038/s41540-017-0045-9 (2018).

15 Davis, M. M., Tato, C. M. & Furman, D. Systems immunology: just getting started. Nature Immunology 18, 725–732, doi:10.1038/ni.3768 (2017).

16 Fagherazzi, G. et al. Protocol for a prospective, longitudinal cohort of people with COVID-19 and their household members to study factors associated with disease severity: the Predi-COVID study. BMJ Open 10, e041834, doi:10.1136/bmjopen-2020-041834 (2020).

17 Tan, L. et al. Lymphopenia predicts disease severity of COVID-19: a descriptive and predictive study. Signal Transduction and Targeted Therapy 5, 33, doi:10.1038/s41392-020-0148-4 (2020).

18 Wajnberg, A. et al. Robust neutralizing antibodies to SARS-CoV-2 infection persist for months. Science 370, 1227–1230, doi:10.1126/science.abd7728 (2020).

19 Takahashi, N. et al. Impaired CD4 and CD8 effector function and decreased memory T cell populations in ICOS-deficient patients. J Immunol 182, 5515–5527, doi:10.4049/jimmunol.0803256 (2009).

20 Bertram, E. M. et al. Role of ICOS versus CD28 in antiviral immunity. Eur J Immunol 32, 3376–3385, doi:10.1002/1521-4141(200212)32:12<3376::AID-IMMU3376>3.0.CO;2-Y (2002).

21 Rha, M. S. et al. PD-1-Expressing SARS-CoV-2-Specific CD8(+) T Cells Are Not Exhausted, but Functional in Patients with COVID-19. Immunity 54, 44–52 e43, doi:10.1016/j.immuni.2020.12.002 (2021).

22 Picozza, M., Battistini, L. & Borsellino, G. Mononuclear phagocytes and marker modulation: when CD16 disappears, CD38 takes the stage. Blood 122, 456–457, doi:10.1182/blood-2013-05-500058 (2013).

23 Villasenor-Cardoso, M. I., Frausto-Del-Rio, D. A. & Ortega, E. Aminopeptidase N (CD13) is involved in phagocytic processes in human dendritic cells and macrophages. Biomed Res Int 2013, 562984, doi:10.1155/2013/562984 (2013).

24 Hadjadj, J. et al. Impaired type I interferon activity and inflammatory responses in severe COVID-19 patients. Science, doi:10.1126/science.abc6027 (2020).

25 Naylor, K. et al. The influence of age on T cell generation and TCR diversity. J Immunol 174, 7446–7452, doi:10.4049/jimmunol.174.11.7446 (2005).

26 Yager, E. J. et al. Age-associated decline in T cell repertoire diversity leads to holes in the repertoire and impaired immunity to influenza virus. J Exp Med 205, 711–723, doi:10.1084/jem.20071140 (2008).

27 Chen, Z. & John Wherry, E. T cell responses in patients with COVID-19. Nature Reviews Immunology 20, 529–536, doi:10.1038/s41577-020-0402-6 (2020).

28 Gittelman, R. M. et al. Diagnosis and Tracking of SARS-CoV-2 Infection By T-Cell Receptor Sequencing. medRxiv, 2020.2011.2009.20228023, doi:10.1101/2020.11.09.20228023 (2021).

29 Snyder, T. M. et al. Magnitude and Dynamics of the T-Cell Response to SARS-CoV-2 Infection at Both Individual and Population Levels. medRxiv, doi:10.1101/2020.07.31.20165647 (2020).

30 Wilmes, P. et al. SARS-CoV-2 transmission risk from asymptomatic carriers: Results from a mass screening programme in Luxembourg. The Lancet Regional Health - Europe 4, 100056, doi:https://doi.org/10.1016/j.lanepe.2021.100056 (2021).

31 Minervina, A. A. et al. Longitudinal high-throughput TCR repertoire profiling reveals the dynamics of T-cell memory formation after mild COVID-19 infection. eLife 10, e63502, doi:10.7554/eLife.63502 (2021).

32 Szabo, P. A. et al. Longitudinal profiling of respiratory and systemic immune responses reveals myeloid cell-driven lung inflammation in severe COVID-19. Immunity 54, 797–814.e796, doi:10.1016/j.immuni.2021.03.005 (2021).

33 Filbin, M. R. et al. Longitudinal proteomic analysis of severe COVID-19 reveals survival-associated signatures, tissue-specific cell death, and cell-cell interactions. Cell Reports Medicine 2, doi:10.1016/j.xcrm.2021.100287 (2021).

34 Liu, C. et al. Time-resolved systems immunology reveals a late juncture linked to fatal COVID-19. Cell 184, 1836–1857.e1822, doi:10.1016/j.cell.2021.02.018 (2021).

35 Bastard, P. et al. Autoantibodies against type I IFNs in patients with life-threatening COVID-19. Science 370, eabd4585, doi:10.1126/science.abd4585 (2020).

36 Acharya, D., Liu, G. & Gack, M. U. Dysregulation of type I interferon responses in COVID-19. Nat Rev Immunol 20, 397–398, doi:10.1038/s41577-020-0346-x (2020).

37 Asselin-Paturel, C. & Trinchieri, G. Production of type I interferons: plasmacytoid dendritic cells and beyond. J Exp Med 202, 461–465, doi:10.1084/jem.20051395 (2005).

38 Vinh, D. C. et al. Harnessing Type I IFN Immunity Against SARS-CoV-2 with Early Administration of IFN-beta. J Clin Immunol, doi:10.1007/s10875-021-01068-6 (2021).

39 Jamieson, T. et al. The chemokine receptor D6 limits the inflammatory response in vivo. Nature Immunology 6, 403–411, doi:10.1038/ni1182 (2005).

40 Chevigné, A. et al. CXCL10 Is an Agonist of the CC Family Chemokine Scavenger Receptor ACKR2/D6. Cancers 13, 1054 (2021).

41 Liu, M. et al. CXCL10/IP-10 in infectious diseases pathogenesis and potential therapeutic implications. Cytokine Growth Factor Rev 22, 121–130, doi:10.1016/j.cytogfr.2011.06.001 (2011).

42 Wong, C. K. et al. Plasma inflammatory cytokines and chemokines in severe acute respiratory syndrome. Clin Exp Immunol 136, 95–103, doi:10.1111/j.1365-2249.2004.02415.x (2004).

43 Yang, Y. et al. Plasma IP-10 and MCP-3 levels are highly associated with disease severity and predict the progression of COVID-19. Journal of Allergy and Clinical Immunology 146, 119–127.e114, doi:10.1016/j.jaci.2020.04.027 (2020).

44 Medford, A. R. & Millar, A. B. Vascular endothelial growth factor (VEGF) in acute lung injury (ALI) and acute respiratory distress syndrome (ARDS): paradox or paradigm? Thorax 61, 621–626, doi:10.1136/thx.2005.040204 (2006).

45 Ray, P. R. et al. A pharmacological interactome between COVID-19 patient samples and human sensory neurons reveals potential drivers of neurogenic pulmonary dysfunction. Brain, Behavior, and Immunity 89, 559–568, doi:https://doi.org/10.1016/j.bbi.2020.05.078 (2020).

46 Xu, Z.-S. et al. Temporal profiling of plasma cytokines, chemokines and growth factors from mild, severe and fatal COVID-19 patients. Signal Transduction and Targeted Therapy 5, 100, doi:10.1038/s41392-020-0211-1 (2020).

47 Chen, Y. G. (https://ClinicalTrials.gov/show/NCT04275414, 2020).

48 Blanco-Melo, D. et al. Imbalanced Host Response to SARS-CoV-2 Drives Development of COVID-19. Cell 181, 1036–1045 e1039, doi:10.1016/j.cell.2020.04.026 (2020).

49 Peyneau, M. et al. Innate immune deficiencies in patients with COVID-19. medRxiv, 2021.2003.2029.21254560, doi:10.1101/2021.03.29.21254560 (2021).

50 Painter, M. M. et al. Rapid induction of antigen-specific CD4+ T cells is associated with coordinated humoral and cellular immune responses to SARS-CoV-2 mRNA vaccination. Immunity, doi:https://doi.org/10.1016/j.immuni.2021.08.001 (2021).

51 Loske, J. et al. Pre-activated antiviral innate immunity in the upper airways controls early SARS-CoV-2 infection in children. Nature Biotechnology, doi:10.1038/s41587-021-01037-9 (2021).

52 Capelle, C. M. et al. Standard Peripheral Blood Mononuclear Cell Cryopreservation Selectively Decreases Detection of Nine Clinically Relevant T Cell Markers. ImmunoHorizons 5, 711–720, doi:10.4049/immunohorizons.2100049 (2021).

53 Robins, H. S. et al. Comprehensive assessment of T-cell receptor beta-chain diversity in alphabeta T cells. Blood 114, 4099–4107, doi:10.1182/blood-2009-04-217604 (2009).

